# Quantifying the dynamics of COVID-19 burden and impact of interventions in Java, Indonesia

**DOI:** 10.1101/2020.10.02.20198663

**Authors:** Bimandra A Djaafara, Charles Whittaker, Oliver J Watson, Robert Verity, Nicholas F Brazeau, Widyastuti Widyastuti, Dwi Oktavia, Verry Adrian, Ngabila Salama, Sangeeta Bhatia, Pierre Nouvellet, Ellie Sherrard-Smith, Thomas S Churcher, Henry Surendra, Rosa N Lina, Lenny L Ekawati, Karina D Lestari, Adhi Andrianto, Guy Thwaites, J Kevin Baird, Azra C Ghani, Iqbal RF Elyazar, Patrick GT Walker

## Abstract

**Background:** As in many countries, quantifying COVID-19 spread in Indonesia remains challenging due to testing limitations. In Java, non-pharmaceutical interventions (NPIs) were implemented throughout 2020. However, as a vaccination campaign launches, cases and deaths are rising across the island.

**Methods:** We used modelling to explore the extent to which data on burials in Jakarta using strict COVID-19 protocols (C19P) provide additional insight into the transmissibility of the disease, epidemic trajectory, and the impact of NPIs. We assess how implementation of NPIs in early 2021 will shape the epidemic during the period of likely vaccine roll-out.

**Results:** C19P burial data in Jakarta suggest a death toll approximately 3.3 times higher than reported. Transmission estimates using these data suggest earlier, larger, and more sustained impact of NPIs. Measures to reduce sub-national spread, particularly during Ramadan, substantially mitigated spread to more vulnerable rural areas. Given current trajectory, daily cases and deaths are likely to increase in most regions as the vaccine is rolled-out. Transmission may peak in early 2021 in Jakarta if current levels of control are maintained. However, relaxation of control measures is likely to lead to a subsequent resurgence in the absence of an effective vaccination campaign.

**Conclusion:** Syndromic measures of mortality provide a more complete picture of COVID-19 severity upon which to base decision-making. The high potential impact of the vaccine in Java is attributable to reductions in transmission to date and dependent on these being maintained. Increases in control in the relatively short-term will likely yield large, synergistic increases in vaccine impact.

**Key questions:** *What is already known?:*

- In many settings, limited SARS-CoV-2 testing makes it difficult to estimate the true trajectory and associated burden of the virus.
- Non-pharmaceutical interventions (NPIs) are key tools to mitigate SARS-CoV-2 transmission.
- Vaccines show promise but effectiveness depends upon prioritization strategies, roll-out and uptake.

*What are the new findings?:*

- This study gives evidence of the value of syndrome-based mortality as a metric, which is less dependent upon testing capacity with which to estimate transmission trends and evaluate intervention impact.
- NPIs implemented in Java earlier in the pandemic have substantially slowed the course of the epidemic with movement restrictions during Ramadan preventing spread to more vulnerable rural populations.
- Population-level immunity remains below proposed herd-immunity thresholds for the virus, though it is likely substantially higher in Jakarta.

*What do the new findings imply?:*

- Given current levels of control, upwards trends in deaths are likely to continue in many provinces while the vaccine is scheduled to be rolled out. A key exception is Jakarta where population-level immunity may increase to a level where the epidemic begins to decline before the vaccine campaign has reached high coverage.
- Further relaxation of measures would lead to more rapidly progressing epidemics, depleting the eventual incremental effectiveness of the vaccine. Maintaining adherence to control measures in Jakarta may be particularly challenging if the epidemic enters a decline phase but will remain necessary to prevent a subsequent large wave. Elsewhere, higher levels of control with NPIs are likely to yield high synergistic vaccine impact.

## INTRODUCTION

As of 3^rd^ February 2021, Indonesia has reported the highest number of confirmed COVID-19 cases (1,111,671) and deaths (30,770) among Southeast Asian countries.[1] Cases were first reported in West Java province, on the island of Java, on 2^nd^ March 2020, amid concern that the disease had circulated widely before.[2,3] The city of Jakarta (the capital of Indonesia) subsequently became the epicenter of the country’s epidemic, following which the disease spread throughout the island.

Non-pharmaceutical interventions (NPIs) have included national social distancing measures encouraging people to work, study and worship at home (15^th^ March)[4]; mandated social distancing measures implemented on 10^th^ April as part of a lockdown, named *Pembatasan Sosial Berskala Besar* or PSBB in Indonesian;[4] and a ban on domestic travel during the month of Ramadan (24^th^ April to 7^th^ June).[5] In June, Indonesia entered the *Adaptasi Kebiasaan Baru* (AKB or ‘new normal’) period where some restrictions were lifted (online supplementary figure S1A; 1B).[4]

During this AKB period, the reported incidence of COVID-19 cases and deaths increased across Indonesia with community transmission evident across the six provinces of Java (online supplementary figure 1C; 1D). PSBB was subsequently reimposed in mid-September for four weeks in Jakarta in response to pressures on healthcare facilities across the city.[6] Cases and deaths continued to rise in 2021, prompting further restrictions in districts across the island from 11^th^ January.[7] On 13^th^ January 2021, Indonesia initiated a nationwide vaccination campaign.[7,8]

**Figure 1.**
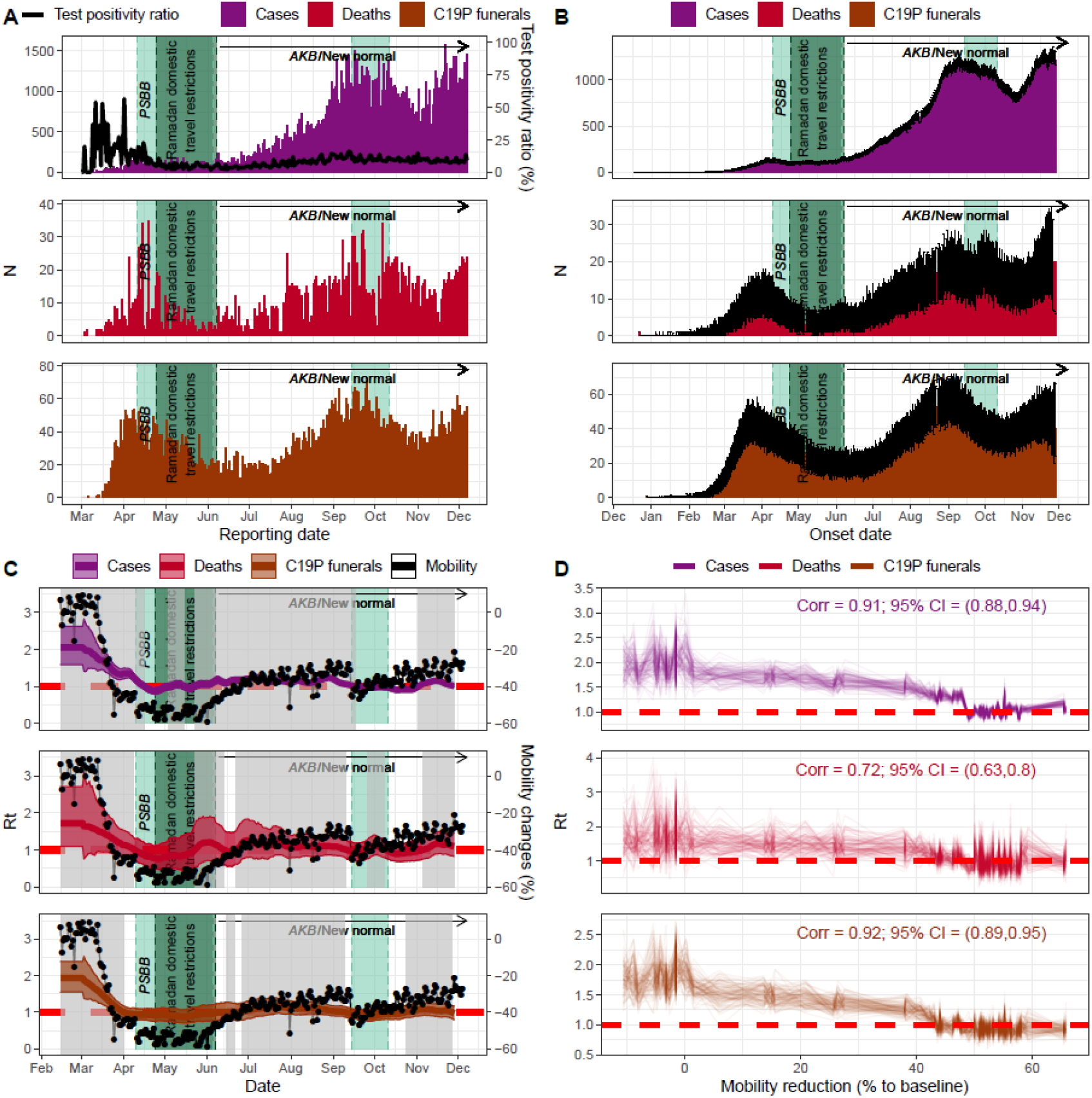
COVID-19-related data, estimated effective reproduction number (*R*_*t*_), and its relationship with the average non-residential mobility changes in Jakarta (epidemiological data up to 7^th^ December 2020; Google Mobility Reports data up to 7^th^ December 2020). A) Daily reported cases, deaths, and C19P funerals in Jakarta. Black line denotes the daily test positivity ratio; B) Reconstructed daily reported cases, deaths, and C19P funerals to reflect the estimated onset date of each observation; C) Coloured lines and regions show, respectively, median and 95% CrI of estimated *R*_*t*_ (left axis) based on the reconstructed data (cases, deaths or C19P funerals). Grey areas denote periods where the estimated median *R*_*t*_ is above 1. Black lines and dots denote average changes in non-residential mobility (right axis); D) The relationship and correlation coefficient between the estimated *R*_*t*_ and the average non-residential mobility reduction (up to 4^th^ June 2020 or before the AKB).

Understanding the trajectory of the epidemic in Java has been challenging. As in many countries,[9,10] testing constraints in Indonesia have limited the extent to which officially confirmed cases reflect underlying trends. Similar concerns exist for mortality data, based upon the high numbers of individuals exhibiting COVID-19 like symptoms who die before receiving a diagnosis.[11,12] In Jakarta, such individuals are buried under strict COVID-19 protocols (C19P). Here we use mathematical modelling approaches incorporating these data, and other measures of suspected mortality, to better understand the dynamics and burden of the epidemic experienced across Java to date, evaluate the impact of control measures, and understand how these past actions will shape future burden and vaccine impact.

## METHODS

### Assessing SARS-CoV-2 transmissibility over time in Jakarta

Daily numbers of confirmed COVID-19 cases, deaths, and C19P funerals[13] were used to reconstruct daily incidence of symptom onset, using delay distributions between symptom onset and case reporting or death derived from individual patient data obtained from the Jakarta Department of Health (online supplementary figure S2). For each data source (cases, deaths, and C19P funerals), 100 reconstructed time-series of daily incidence of symptom onset were generated, with estimates also adjusted for right-censoring in individuals where outcomes had yet to occur (online supplementary methods section S2).

These reconstructed time series were translated into estimates of the daily effective reproduction number (*R*_*t,case*_ for cases, *R*_*t,death*_ for deaths, and *R*_*t,funeral*_ for funerals) in Jakarta using EpiEstim.[14,15] This package estimates *R*_*t*_ using a branching process-based estimator that incorporates information on the serial interval distribution and dates of onsets of symptoms. Correlations between estimated *R*_*t*_ and the average daily changes in non-residential mobility[16] were assessed based on 1,000 posterior samples from each estimated *R*_*t*_ time series and compared using Pearson’s correlation coefficient formula.

### Modelling subnational COVID-19 spread across Java

We developed a district-level metapopulation model to explore the expected spread of COVID-19 across the island of Java (online supplementary methods section S5). For each district, stochastic differential equations representing a Susceptible-Exposed-Infected-Recovered (SEIR) model were implemented. Movement matrices were derived from anonymized mobile phone data, with separate matrices calculated for the high-migration period of Ramadan. Disease severity parameters were adjusted to account for the demography of each district. Transmissibility of the virus over time was calculated under the assumption that the relationship between mobility and *R*_*t*_ observed in Jakarta was informative across the rest of the island, exploring multiple assumptions about the transmissibility of COVID-19 in rural districts relative to urban districts (online supplementary table S3).

We simulated five different scenarios to assess the impact of restrictions earlier in the pandemic in Indonesia (namely PSBB and Ramadan movement restrictions) on COVID-19 deaths and hospitalisation rates across Java (online supplementary table S4). The baseline scenario simulated an epidemic across Java based on the observed mobility patterns and an assumption that longer-distance between-district movement was reduced to a greater extent than within-district (a further reduction of an odds ratio (OR) of two relative to pre-pandemic levels) and severely curtailed (a 95% reduction) during Ramadan. Counter-factual scenarios for the absence of restrictions during Ramadan, assuming no movement restrictions between-district in place, were also simulated. Uncertainty in the incremental effect of the absence of between-district movement restrictions during Ramadan on the population-level transmission was then captured in three scenarios: a) Ramadan 1, that they had no impact (i.e., within-district movement would remain the same as the baseline scenario in the absence of Ramadan-specific intervention); b) Ramadan 2, they were responsible for 75% of the reduction in *R*_*t*_ relative to *R*_0_; c) Ramadan 3, without restrictions, transmission would have returned to *R*_0_ levels during Ramadan. Lastly, the unmitigated scenario simulated an epidemic across Java without movement restrictions and without reductions in transmissibility due to behaviour change and/or control measures.

### Assessing the current province-level spread of the pandemic in Java and generating future scenarios

To estimate the recent trajectory of the epidemic and current cumulative levels of spread within each province, we adapted an existing modelling framework allowing the relationship between mobility and transmission to vary over time.[9] This allows us to capture the observed decoupling between aggregated movement patterns and burden in the ‘new normal’ period and simulate scenarios of future spread within each province. We fit this modelling framework both to officially reported COVID-19 deaths, as well as estimated suspected deaths, which include deaths of probable cases (i.e., patients with clinical criteria or chest imaging suggestive of COVID-19), which have been published by WHO Indonesia.[4] As the published suspected deaths data are only available on a weekly basis between 1^st^ June to 29^th^ November 2020, we augmented the data to reflect the entire time-period of the epidemic based upon the proportion of all suspected deaths (i.e., probable and confirmed) that were confirmed by each province in the period covered by the WHO situation reports (online supplementary methods section S6).

Our future scenarios are projected based on a future ‘reproduction number under control’, *R*_*c*_, defined similarly to *R*_0_ as the average number of secondary infections within an entirely susceptible population but incorporating the impact of NPIs (and, equivalently, *R*_*t*_ but not incorporating the effects of population-level immunity such that *R*_0_ > *R*_*c*_ > *R*_*t*_). We evaluated three scenarios: a ‘current trajectory’ scenario (where the current trajectory of the epidemic continues with approximated *R*_*c*_ = 1.25), a ‘suppression’ scenario (where the transmission in the population is assumed to be immediately suppressed with *R*_*c*_ = 0.75) and an ‘unmitigated’ scenario (where the epidemic was assumed to be uncontrolled with *R*_*c*_ = 2.00).

Our first set of projections were generated from 2^nd^ September 2020 onwards.[17] At that time, policymakers were attempting to understand the potential benefits of the implementation of further NPIs, such as the reimposition of PSBB in Jakarta, which was then scheduled to be implemented on 14^th^ September,[6] in the context of no vaccine yet being any available. These scenarios evaluated the potential trajectory of the epidemic throughout 2021, including the impact of a ‘return-to-normal’ (*R*_*c*_ = 2.00) once burden had returned to low-levels (median of simulated trajectories reached less than 7 cumulative deaths over 7 days period).

Our current set of projections are generated from 7^th^ December and in the context of an imminent vaccine campaign. Given the large remaining uncertainties in roll-out and effectiveness, we do not incorporate any role of the vaccine. Instead, we aim to understand how different scenarios involving NPIs over the next few weeks and months will shape the potential longer-term effectiveness of future strategies in which vaccines will likely feature as a major component. To do this, we evaluate how both the number of lives lost, and number of lives that remain to be saved is likely to change incrementally by month according to the same future scenarios (i.e., current mitigation, suppression and unmitigated), relative to an unmitigated epidemic from the date of our projection (7^th^ December 2020).

## RESULTS

### Understanding initial establishment, transmission, and dynamics of SARS-CoV-2 in Jakarta

Figure 1A shows the daily reported cases, deaths, test positivity ratios, and funerals with C19P in Jakarta, transformed into inferred dates of symptom onset using the relevant delay distributions (figure 1B). We estimate that 31 (22-41 95% CrI) and 124 (107-139 95% CrI) confirmed deaths and C19P funerals (assuming all funerals represent deaths due to COVID-19) had symptom onset occurring before 2^nd^ March when COVID-19 was first identified in Indonesia. We estimate 10,950 (7,530-14,040 95% CrI) infections based on confirmed deaths or 42,100 (36,280-47,570 95% CrI) based on C19P funerals (reflecting an assumption that all undiagnosed individuals with a C19P funeral would have tested positive) had occurred in Jakarta by 2^nd^ March.

Reported cases in Jakarta appear to indicate two epidemic peaks to date (around mid-April and mid-September, when PSBB was imposed), with the number of cases reported during the second peak far higher than the first (figure 1B). However, the test-positivity rate declined in the first half of 2020, indicating increased testing rates and case-ascertainment, which complicates the interpretation of trends based on case data alone. Indeed, data on C19P funerals suggest that the peak in infections likely occurred in mid-March and that infection levels during the second peak were at levels comparable to their initial peak.

Our branching-process-based estimates of *R*_*t*_ support the substantial impact of NPIs when applied to all three metrics. We estimate *R*_*t*_ to be between 1.5 and 2.5 initially, subsequently declining to below 1 during the first PSBB period, followed by a more recent increase to slightly above 1 as Jakarta entered the transitional PSBB in early June. The reimposition of the second PSBB in September also brought the *R*_*t*_ to below 1 (figure 1C). Before the lifting of the first PSBB, *R*_*t*_ estimates show a strong and significant correlation (0.91, 0.72, and 0.92 for cases, deaths, and C19P funerals, respectively, all with p<0.001) with observed mobility patterns as measured by Google Mobility Reports (figure 1D). Estimates based upon funeral trends support a more rapid, larger, and more sustained impact of interventions than those based upon case-reporting. The correlation with within-city mobility is lowest for the deaths data, where substantial variation in day-to-day death reporting leads to more unstable *R*_*t*_ estimates over time. Calculating the correlation between mobility and *R*_*t*_ before and after the AKB period suggests a decoupling between transmission and mobility, whereby estimates of *R*_*t*_ during periods of equivalent levels of mobility during AKB are lower than estimates obtained before AKB (online supplementary figure S4).

### Understanding COVID-19 risk and subnational spread of SARS-CoV-2 across Java

Substantial variations exist across the island in terms of demography, healthcare capacity, and between-district mobility. The proportion of individuals over the age of 50 is typically higher (26%) in rural districts than urban ones (19%) (figure 2A). There are also substantial disparities in healthcare availability, ranging from the comparatively well-resourced Jakarta setting (2.22 hospital beds per thousand population) to the poorer, more rural setting of Tasikmalaya in West Java (0.18 hospital beds per thousand population) (figure 2B). Patterns of between-district mobility outside of the window of the pandemic, estimated using mobile phone data over the period of 1^st^ May 2011 - 30^th^ April 2012, highlight the extent to which these settings are connected. Between-district connectivity is particularly high during the Ramadan period, with large-scale movements from densely populated Jakarta to other more rural regions with lower availability of healthcare (figure 2C; 2D). Applying our modelled relationship between mobility and *R*_*t*_ obtained from the Jakarta C19P funeral data (figure 2E) to trends in mobility data from the remaining provinces in Java suggests large reductions in transmission in all provinces coinciding with the PSBB period. However, they also suggest that measures were sufficient to bring *R*_*t*_ below 1 for a sustained period only in Jakarta and Yogyakarta. Increases in mobility occurred either during early May (Banten, West Java, Central Java, and East Java) or alongside the establishment of the AKB in June (Jakarta and Yogyakarta), leading to corresponding increases in our estimates of *R*_*t*_ (figure 2F).

**Figure 2.**
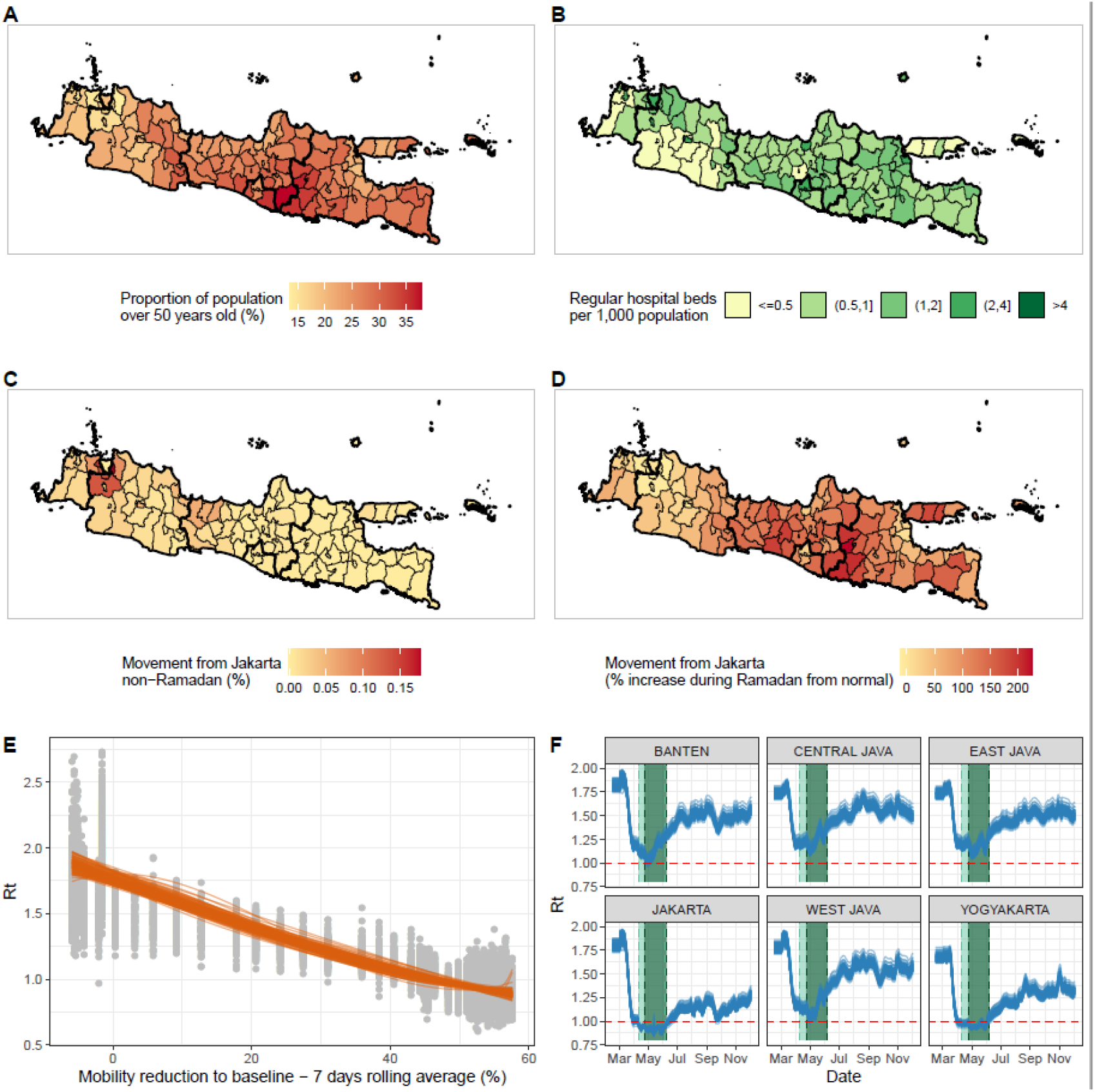
Key factors that are affecting the spread and severity of COVID-19 epidemic in Java, Indonesia. A) Proportion of the population aged over 50 years old at the district level; B) Number of regular hospital beds per one thousand population at the district level; C) Proportion of Jakarta residents who spent their day in other districts in Java during a non-Ramadan period; D) Increased proportion of people of Jakarta who spent their day in other districts in Java during Ramadan compared to the non-Ramadan period; E) The relationship between the estimated *R*_*t*_ values based on C19P funerals data and average reduction in non-residential mobility in Jakarta. Orange lines show the modelled smoothing spline relationship for 100 samples; and F) Extrapolations of *R*_*t*_ values in provinces in Java based upon Google Mobility trends for each province and the 100 sampled smoothing splines in E.

These estimates were integrated into our meta-population model (figure 3A). Estimates of deaths in the baseline scenario were consistent with observed qualitative patterns prior to the shift to the AKB phase of the epidemic in early June. The epicenter shifted over time from Jakarta to satellite towns and other provincial capitals, and with Yogyakarta remaining least affected. Our baseline scenario’s median deaths fall within the range of cumulative confirmed and suspected deaths up to 31^st^ May 2020 and the number of confirmed and suspected deaths between 13^th^-31^st^ May 2020 in most provinces (table 1). Total suspected deaths fell within the model’s uncertainty bounds for most provinces except Jakarta and Central Java (table 1).

**Table 1.** Total number of estimated deaths based on model simulations of the actual epidemic scenario and its counterfactual of an unmitigated scenario (assuming no interventions) from the beginning of the epidemic up to 31^st^ May 2020. Values inside the brackets denote 95 percentile range of simulations. Suspected deaths are a combination of confirmed and probable COVID-19 deaths.

| Province | Confirmed deaths 13<sup>th</sup>-31<sup>st</sup> May (WHO Indonesia situation report 10[4]) | Suspected deaths 13<sup>th</sup>-31<sup>st</sup> May (WHO Indonesia situation report 10[4]) | Baseline model scenario deaths 13<sup>th</sup>-31<sup>st</sup> May | Confirmed deaths up to 31<sup>st</sup> May[13,30] | Suspected deaths up to 31<sup>st</sup> May (provincial data collated by KawalCOVID19 [31]) | Baseline model scenario deaths up to 31<sup>st</sup> May | Unmitigated counterfactual deaths up to 31<sup>st</sup> May | Averted deaths up to 31<sup>st</sup> May (unmitigated – baseline) |
| --- | --- | --- | --- | --- | --- | --- | --- | --- |
| Jakarta | 74 | 447 | 158 (47-333) | 520 | 2,435 | 810 (292-1777) | 16,356 (7,896-21,593) | 15,560 (7,567-19,691) |
| West Java | 46 | 351 | 197 (55-525) | 135 | 653 | 525 (149-1,368) | 19,733 (5,682-39,876) | 19,151 (5,516-38,400) |
| Central Java | 4 | 269 | 88 (36-216) | 66 | 666 | 203 (67-511) | 6,321 (2,147-16,052) | 6,068 (2,056-15,485) |
| Yogyakarta | 0 | 1 | 2 (0-8) | 9 | 29 | 6 (1-33) | 401 (138-1,097) | 397 (132-1,088) |
| East Java | 241 | 458 | 437 (98-944) | 395 | 1,127 | 1,091 (226-2,646) | 12,182 (3,625-20,800) | 10,997 (3,277-27,045) |
| Banten | 13 | 47 | 82 (20-224) | 67 | 332 | 229 (66-711) | 7,302 (2,180-14,732) | 7,0759 (2,111-14,141) |
| Java island total | 378 | 1,638 | 983 (360-1,930) | 1,192 | 5,242 | 2,912 (1,109-5,851) | 59,896 (26,787-112,795) | 57,030 (24,843-105,378) |

**Figure 3.**
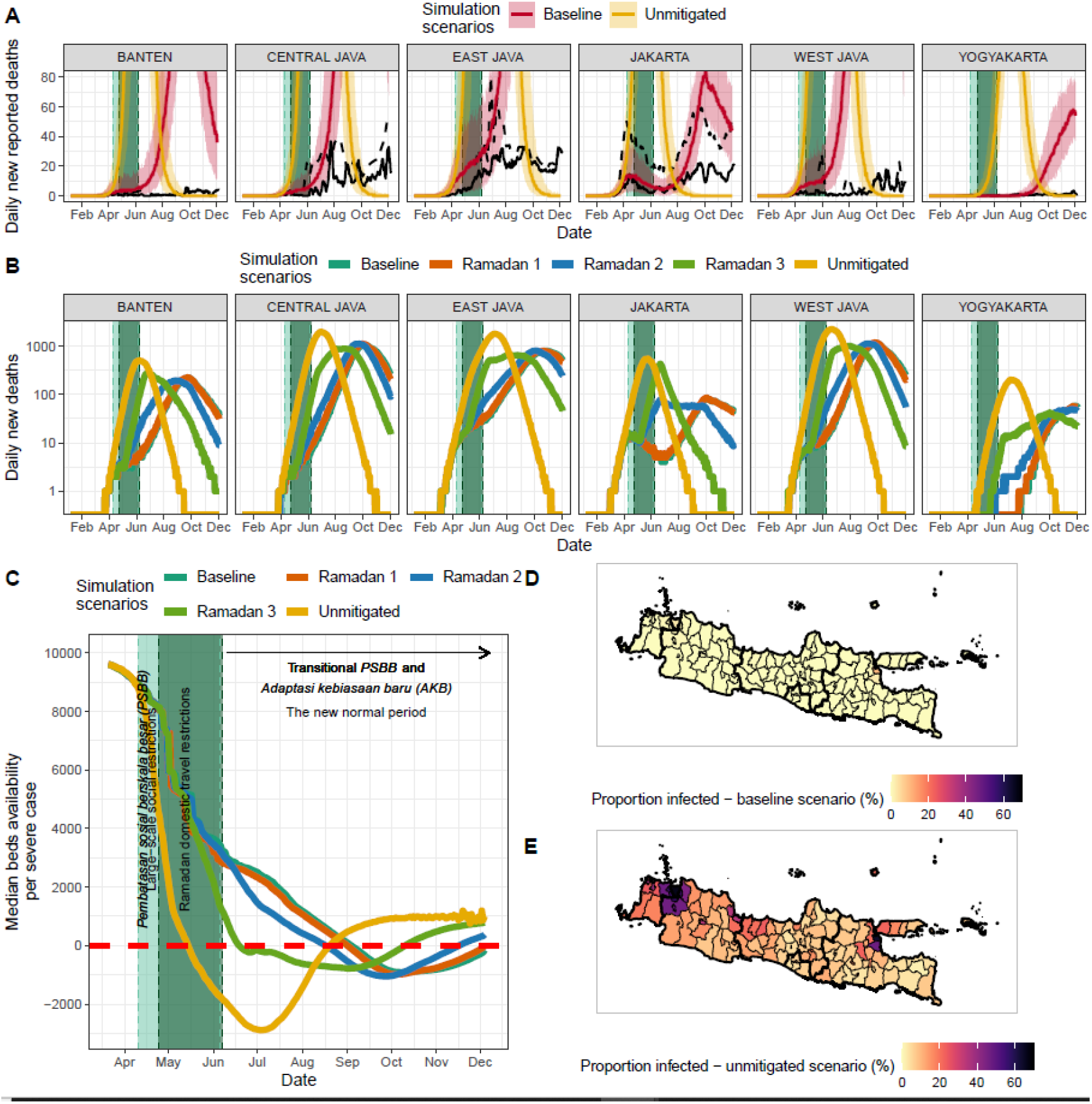
Metapopulation model simulation results. A) Comparison of model simulations in the baseline scenario (red lines and their shaded 95% uncertainties ranges) and unmitigated scenario (yellow lines and their shaded 95% uncertainties ranges), and daily confirmed (solid black lines) and suspected (dashed black lines) deaths from COVID-19; B) Model simulations in five different scenarios: 1) Baseline scenario as shown in A, 2) Ramadan counter-factual 1 where it is assumed that there is no movement restrictions during the Ramadan period and *R*_*t*_ values are similar to the baseline scenario, 3) Ramadan counter-factual 2 where it is assumed that there is no movement restrictions during the Ramadan period and *R*_*t*_ values are 75% of each district’s *R*_0_ value, 4) Ramadan counter-factual 3 where it is assumed that there are no movement restrictions during the Ramadan period and *R*_*t*_ values are each district’s *R*_0_ value and 5) Unmitigated scenario where no interventions since the beginning of the epidemic are assumed; C) Median hospital beds availability per severe COVID-19 case over time based on different simulation scenarios; D) Proportion of people infected based on the actual scenario up to 31^st^ May 2020 (before AKB/the ‘new normal’) at the district level; and E) Proportions of people infected based on the unmitigated scenario up to 31^st^ May 2020 (before AKB) at the district level.

The scenarios estimates are consistent with reductions in contact rates serving to reduce spread, reduce healthcare demand and avert mortality prior to AKB phase: an estimated 57,000 (24,800-105,400, 95% UI) deaths averted when compared to an effectively unmitigated epidemic with *R*_*t*_ = 2 throughout this period (which we estimate would have resulted in 59,900 (26,800-112,800, 95% UI deaths). These numbers do not consider the effects of healthcare services becoming overwhelmed (as shown by the negative values of the median number of hospital beds available per COVID-19 case needing hospitalisation under the unmitigated epidemic scenario; figure 3C) on both direct and indirect mortality, an impact which would likely have been sizable given the wider spread to more rural settings with more scarce healthcare provision in our unmitigated scenario (figure 3D; 3E).

Our baseline scenario increasingly over-predicts deaths in most provinces during the AKB. This is in line with our results suggesting a decoupling of within-province mobility from virus transmissibility over that period.

### Estimating current COVID-19 burden, modelled future scenarios, and estimated vaccines impact in Java

Our projections generated 2^nd^ September 2020[17] (figure S13) suggested that whilst *R*_*t*_ was well below that observed at the beginning of the epidemic, this was driven primarily by the impact of control measures rather than the accumulation of population-level immunity. As a result, in the absence of additional control measures, death rates were likely to rise for the remainder of the year in all provinces, pushing all provinces beyond available hospital capacity. We found that reimplementation of PSBB could largely prevent capacity from being exceeded but would not prevent a subsequent wave if such control was not maintained.

Subsequently, between our two sets of simulations (2^nd^ September 2020 and 7^th^ December 2020), both confirmed deaths and our inferred estimates of total suspected deaths increased from 5,108 to 11,370 and 12,254 to 26,206, respectively, across Java. At the island level, the estimated attack rates on both time points increased from 1.21% to 2.57% and 2.95% to 6.03% based on confirmed deaths and assuming all suspected deaths as COVID-19 deaths, respectively (figure 4A). At the province level, estimates of attack rate and total burden from COVID-19 differ quite significantly, with Jakarta accumulating the highest attack rates in the region by 7^th^ December 2020 (figure 4B; table S6). In all provinces and based on models fitted to either suspected or confirmed deaths, there were consistent increases of around 2-3 times on the province-level attack rate from 2^nd^ September to 7^th^ December 2020. However, as seen at the island level, discrepancies between the estimated attack rates based on the model fitted to suspected deaths and confirmed deaths data were still observed at the province-level, with the highest difference observed in Jakarta.

**Figure 4.**
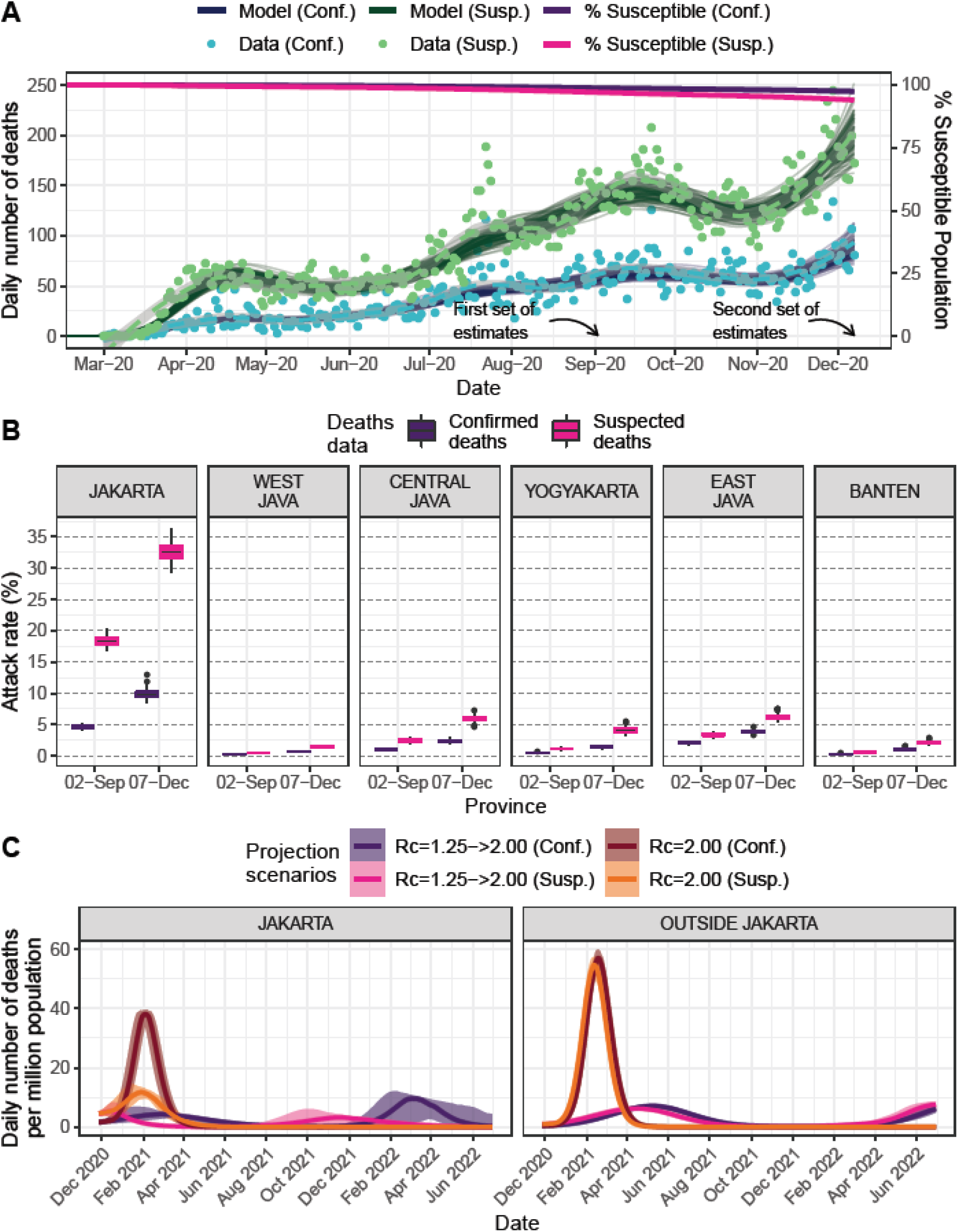
A) Model fitting to confirmed and suspected (both confirmed and probable) COVID-19 related deaths and inferred population susceptibility in Java; Green and blue dots show data on reported and suspected respectively (where suspected includes augmented estimate of probable deaths in provinces outside Jakarta), with associated median (lines) and 95% CrI (shaded areas) of model fits. B) Estimated province-level attack rates (cumulative proportion infected) based on confirmed (purple) and suspected (pink) COVID-19 related deaths. C) Projections of daily number of deaths due to COVID-19 based on four different transmission scenarios.

Projections of future scenarios from December 2020 (figure 4C; and figure S14 for province-level breakdown), incorporating these changes in estimated attack rate and extrapolating current trends of *R*_*c*_, leads to the projected daily incidence of mortality across the island continuing to grow throughout the first half of the 2021 irrespective of whether reported or suspected mortality are more reflective of true direct COVID-19 mortality. In this scenario, with future *R*_*c*_ = 1.25, the epidemic would be projected to peak earliest in Jakarta, driven by the higher degree of population-level immunity implied by the higher cumulative attack rate to date. This peak’s timing is sensitive to the mortality metric the model is calibrated to, with projected peaks occurring early in 2021 for a current scenario based upon suspected deaths and towards the end of the first quarter of 2021 based upon reported deaths. However, in all provinces, at no point in any of our current scenarios was there sufficient population-immunity to preclude a subsequent upsurge in deaths if transmission levels returned to those estimated at the beginning of the pandemic (*R*_*c*_ ≈ 2.00).

Figure 5A shows trajectories of the three different future scenarios summarized in terms of the proportion of lives lost before the beginning of a month (figure 5B) and the total remaining lives to be saved (deaths that can still be averted) after the start of the month (figure 5C). We estimate that reimposing suppression scenarios in areas where epidemics are on an upwards trajectory would significantly reduce lives lost during a period whilst the vaccine is rolled out. In some settings, such as Jakarta, assuming all suspected deaths were COVID-19 deaths, a combination of control measures currently in place and increasing levels of population immunity may combine to reduce transmission and burden to low-levels temporarily. At this point, the future incremental impact of suppression measures would likely be limited. However, in such scenarios, the high loss of life we estimate would occur in a subsequent lifting of control measures, which highlights the need for ongoing control measures and the substantial remaining incremental value of an effective vaccination campaign (figure 5C).

**Figure 5.**
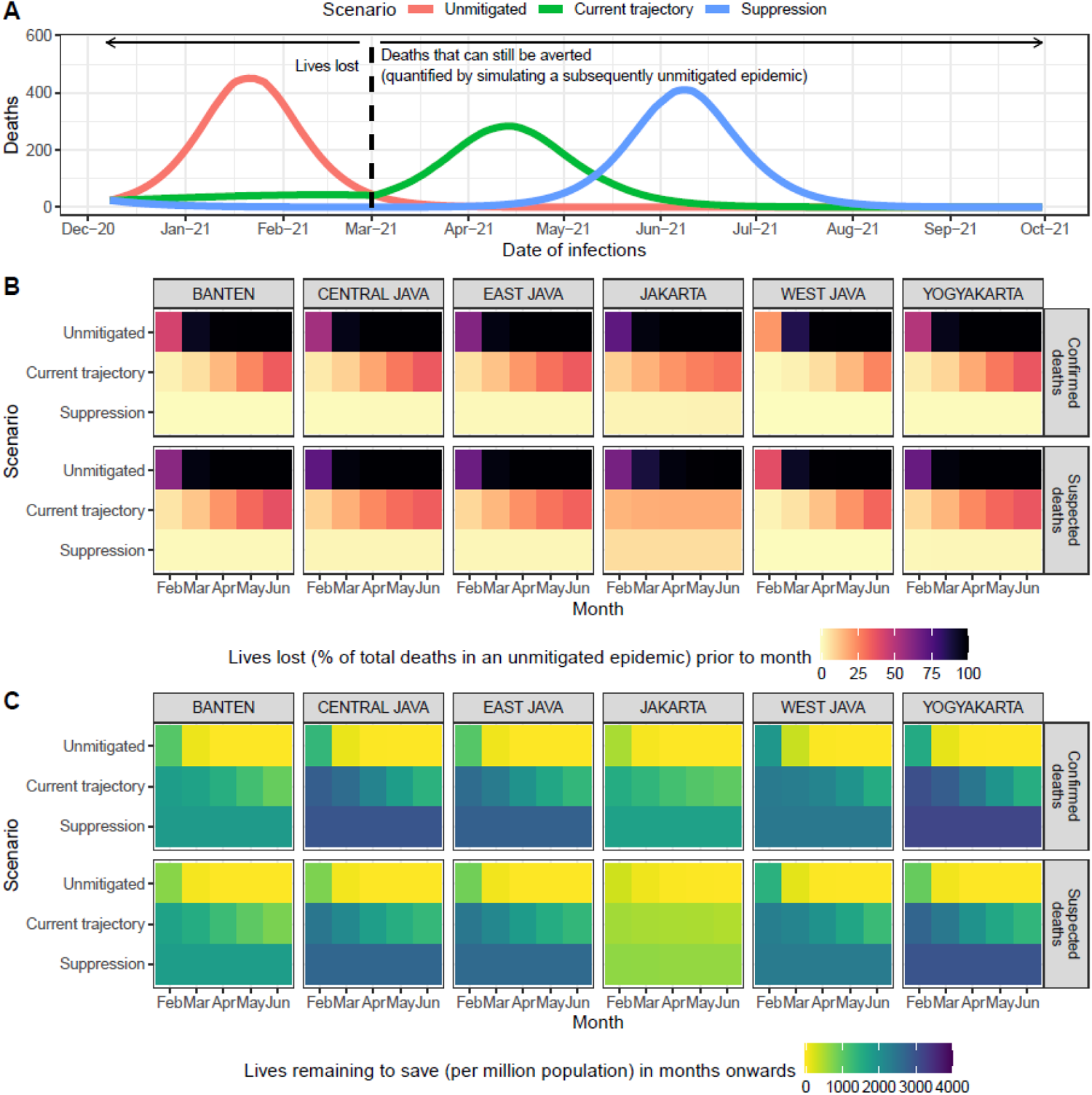
A) An illustration of future scenario projections and how to define the number of lives lost and the number of deaths that can still be averted after a certain time point. The graph shows simulations based on a model fitted to confirmed COVID-19 deaths in Jakarta, which subsequently ‘returning to normal’ on 1^st^ March 2021; B) Projected percentage of lives lost (compared to total deaths from an unmitigated epidemic scenario) prior to the start of each month from February to June 2021, based on each simulation scenario and model fitted to confirmed or suspected deaths in each province in Java; C) Projected number of lives remaining to be saved (or deaths that can still be averted) per million population after the start of each month from February to June 2021, based on each simulation scenario and model fitted to confirmed or suspected deaths in each province in Java.

## DISCUSSION

Our analysis uses C19P funeral data in Jakarta to highlight the considerable benefits of using syndromic measures of COVID-19 mortality for surveillance purposes– particularly for countries such as Indonesia, where testing capacity has been severely strained in the face of the pandemic. C19P funerals and other measures of suspected mortality provide an alternative lens through which to understand COVID-19 burden and dynamics but do not allow precise measurement. Without confirmed diagnoses, the proportion of these individuals who were infected will always be unknown and liable to vary spatiotemporally, as will the extent to which measures of suspected deaths represent all deaths of individuals displaying COVID-19 symptoms. These data also support the substantial circulation of SARS-CoV-2 in Indonesia well before the first confirmed COVID-19 case[3] and the higher impact of the virus than suggested by confirmed deaths alone. Simultaneously, they also indicate an earlier decline in transmission during the early stages of the pandemic, coinciding with reductions in mobility, and more sustained declines in transmissibility in response to NPIs than observed in confirmed deaths, a metric which is likely sensitive to limitations in testing. We also found these effects consistent with NPIs substantially attenuating spread across Java, including to older, more rural populations with lower access to healthcare.

Better quantifying impact in the past helps us to better understand likely scenarios in the future. In our first set of projections in September 2020,[17] we suggested that C19P data could indicate up to a four-fold increase in cumulative exposure to the virus relative to confirmed deaths. However, even when assuming a higher burden of the disease in the population, immunity accumulated at the population-level would not prevent the burden from increasing throughout the remainder of 2020. We also suggested that measures to suppress the virus could prevent such a scenario but would need to be sustained to prevent further upsurges. From early 2021, these projections appear to have been valid as transmission declined in Jakarta whilst PSBB was implemented between 14^th^ September – 11^th^ October 2020 but subsequently resurged once restrictions were lifted. Overall, Java’s current epidemiological situation is substantially worse than in September, with record deaths reported week-on-week.[18]

Despite the qualitative validity of our September projections, there are multiple limitations associated with these analyses that should be noted, particularly as our current estimates of attack rates in all provinces in Java have increased substantially. Firstly, it remains difficult to say what level of population-immunity is required to achieve herd immunity as individual immune responses to the virus are still not yet well understood (including strength and duration),[19,20] and heterogeneity in population mixing beyond age-structure likely play important roles.[21,22] Moreover, our estimates of counterfactual ‘return-to-normal’ scenarios rely upon an estimate of *R*_*c*_ = 2.00 from the early stage of the epidemic in Jakarta, a period in which data were particularly limited and, given increasing global concern around the pandemic, where a degree of behavior change relevant may have already occurred. As this estimate is also below those estimated in the early stage of the epidemic from other settings,[23] this estimate may represent a conservative measure of the basic reproduction number. These limitations around the inherent transmissibility and critical immunity threshold to control the virus need to be further considered in the light of recent concerns of new variants of concern across the globe which appear more transmissible.[24–26] There have also been observed resurgences in populations where attack rates had likely passed many estimates of the herd immunity threshold.[24,27]

The initiation of the vaccination campaign in the middle of January[8] provides hopes for more sustainable control of the virus. However, challenges in access and distribution,[28] uncertainty in vaccine efficacy,[28] and possible introductions of variants able to escape immune response[29] could hamper the life-saving impact of the vaccination program. Despite the unprecedented speed of global vaccine development, our study indicates that in the absence of NPIs implemented over the previous year, this campaign would have been too late to prevent most deaths that currently remain avertable. It also highlights the ongoing value and need to maintain current control measures during the coming months as the vaccine is rolled out. Given low estimated attack rates and current increasing trends in transmission across much of the island, our results suggest that further measures aimed towards suppression of the disease over the next few months would substantially increase the proportion of the population who receive the vaccine prior to being exposed to infection, leading to a likely substantial incremental impact of the vaccination campaign. However, we are not able to capture the socio-economic costs of such approaches, which would need to be factored into balanced decision-making.

The case for maintaining or increasing control measures is likely more intuitive to grasp in circumstances where the incidence of cases and deaths continues to rise. However, our projections for Jakarta, particularly those incorporating suspected deaths, suggest it is plausible that population-level immunity may contribute to a decline in the observed rate of new infections, and subsequently, deaths even in the absence of a vaccine in the coming months. Such an effect may have consequences for the perceptions of both the vaccine’s relative impact (for example, if deaths start to decline as the vaccine is rolled out in Jakarta but not in other provinces) as well as the ongoing need for NPIs and/or high vaccine uptake. In such circumstances, our counterfactual of a ‘return-to-normal’, which produces major upsurges in cases and deaths in every province regardless of mortality metric, provides a valuable reminder that the epidemic, and the need to control it, is far from over in any region of Java.

## Data Availability

Data used are publicly available from these websites:
1. https://corona.jakarta.go.id/id/data-pemantauan
2. https://covid19.go.id/peta-sebaran
3. http://kcov.id/daftarpositif
4. https://www.who.int/indonesia/news/novel-coronavirus/situation-reports

## Contributors

BAD, CW, OJW, ACG, IRFE, and PGTW conceived and designed the study. W, DO, VA, and NS collected, verified, and provided data interpretation. BAD, CW, OJW, RV, NFB, SB, PN, IRFE, and PGTW involved in data analysis and interpretation. BAD, CW, OJW, and PGTW drafted the paper. BAD, CW, OJW, RV, NFB, W, DO, VA, NS, SB, PN, ESS, TSC, HS, RNL, LLE, KDL, AA, GT, JKB, ACG, IRFE, and PGTW critically revised the manuscript for important intellectual content and all authors gave final approval for the version to be published.

## Supplementary Material

### S1. Data Sources and Curation

#### Epidemiological data sources for Jakarta

Epidemiological data for Jakarta were obtained from the official Jakarta COVID-19 data monitoring website (https://corona.jakarta.go.id/id/data-pemantauan).^1^ This data comprises daily reported cases, reported deaths, funerals with COVID-19 protocol (C19P), and the number of tests. We collate the data for analysis up to 7^th^ December 2020.

Anonymous individual data of 11,280 confirmed COVID-19 cases up to 29^th^ June 2020 in Jakarta were obtained from the Jakarta Department of Health. Data consist of dates of onset of symptoms, dates of attendance in the hospital, and dates of deaths. The individual data were used to estimate the delay distributions between onset to diagnosis and onset to death.

#### Epidemiological data sources for five other provinces in Java (Banten, West Java, Central Java, Yogyakarta, and East Java)

Daily reported cases and reported deaths data for five other provinces in Java were obtained from an independently curated online spreadsheet by a crowdsource organisation KawalCOVID19 (kcov.id/daftarpositif)^2^ based on the daily publication by Indonesia COVID-19 National Task Force^3^ (data for analysis were collated up 7^th^ December 2020). The weekly number of deaths of suspected and probable cases was obtained from WHO Indonesia situation reports 13-36.^4^

**Figure S1.**
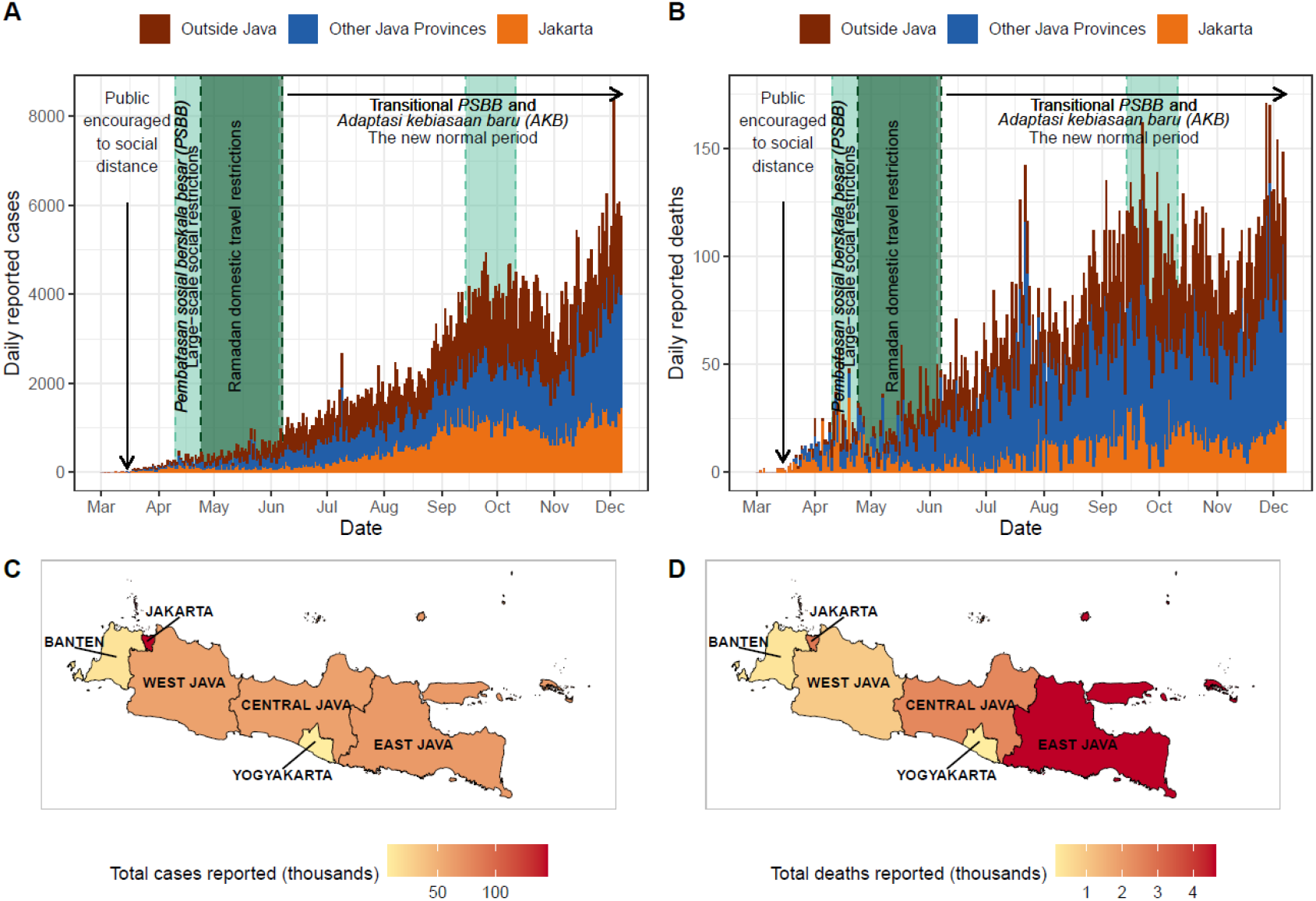
Burden of COVID-19 and timeline of interventions in Indonesia (data up to 7^th^ December 2020). A) Daily number of reported COVID-19 cases; B) Daily number of reported COVID-19 deaths; C) Total reported COVID-19 cases at province level in Java island; and D) Total reported COVID-19 deaths at province level in Java island.

#### Call detail records data

Anonymised call detail records (CDRs) data from one of the biggest telecommunication companies in Indonesia over the period of 1^st^ May 2011 to 30^th^ April 2012 were used to estimate between-district movement matrices for the Ramadan and non-Ramadan period. The CDRs data were collected daily with a total of 266 billion records and 137 million unique SIM cards. There was a total of 17,319 mobile phone towers operated during the period of CDRs data collection across the country.

#### Province-level mobility changes

Province-level mobility changes data were acquired from the freely-available Google COVID-19 Community Mobility Reports (https://www.google.com/covid19/mobility/).^5^ Google Mobility Reports data up to 7^th^ of December 2020 were used for the analysis.

#### Healthcare capacity data

District-level hospital and ICU beds data were obtained from the Online Healthcare Facilities (*Fasyankes Online*) website by the Directorate General of Health Services (*Ditjen Yankes*) of the Ministry of Health of the Republic of Indonesia.^6^

#### Dedicated COVID-19 isolation beds and ICU beds data

Data for the capacity of dedicated COVID-19 isolation beds and ICU beds were obtained from a report from the Ministry of Health in August 2020.^7^

### S2. Reconstruction of frequency of onset

Daily reported cases, reported deaths, and C19P funerals data in Jakarta were reconstructed to represent the onset day of each reported event using estimates of the distribution of delays between onset and diagnosis and onset and death. Each C19P funeral was assumed to occur the day following the date of death.

The distributions of onset to diagnosis and onset to death of confirmed COVID-19 cases were estimated by fitting discretised Gamma distributions^8^ to the individual patient data obtained from the Jakarta Department of Health (**Figure S2**). The mean estimate of the onset to diagnosis delay was 7.62 days, with a standard deviation of 7.51 days. The mean estimate of the onset to death delay was 15.87 days, with a standard deviation of 9.34 days. 100 sets of reconstructed daily frequencies of onset of cases, deaths, and funerals were then calculated on the basis of 100 draws for each reported event from these distributions (respectively onset to hospitalisation, onset to death, and onset to funeral). Adjustment for right censoring occurring due to individuals currently with symptoms but have yet to reach outcome was carried out by dividing the inferred onset frequency on a given day by the cumulative probability it would have been observed by the last date within the dataset.

**Figure S2.**
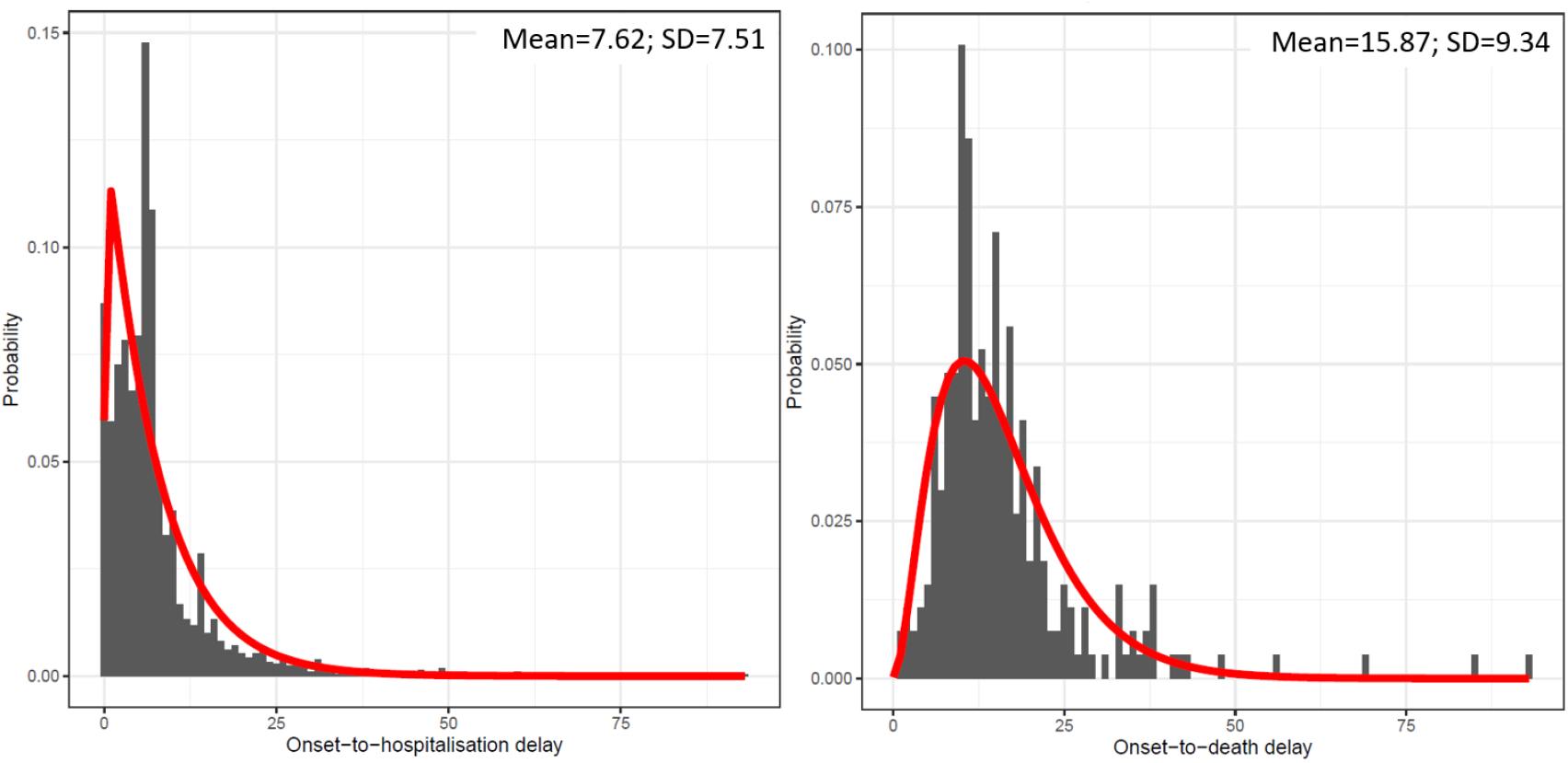
Discretised Gamma distribution fittings to onset-to-hospitalisation delay data (left) and onset-to-death delay data (right).

### S3. Effective reproduction number (*R*_*t*_) calculations based on reconstructed epidemiological data in Jakarta and its relationship with daily mobility changes

The daily effective reproduction number in Jakarta was estimated using the EpiEstim R package^9,10^ for each reconstructed cases (*R*_*t, cases*_), deaths (*R*_*t, deaths*_), funerals (*R*_*t, funerals*_) data. *R*_*t*_ at the beginning of the epidemic was estimated for the period before 2^nd^ March 2020 (the day where the country’s first two cases were announced). Subsequently, *R*_*t*_ was estimated over a weekly sliding window, with a mean and standard deviation of serial interval distribution were assumed to be 6.3 and 4.2 days, respectively.^11^

1,000 random samples were drawn from the posterior samples of the estimated *R*_*t,cases*_, *R*_*t,deaths*_, and *R*_*t,funerals*_ at each timepoint. Pearson’s correlation coefficients for estimates based on cases, deaths, and funerals data were calculated against the daily average changes in non-residential mobility in Jakarta based on Google mobility estimates. The daily average changes in non-residential mobility are the average of changes of retail and recreation, grocery and pharmacy, parks, transit stations, and workplaces types of mobility to each respective baseline.

The 7-day rolling average changes of non-residential mobility were fitted to 100 posterior samples of the estimated *R*_*t,funerals*_ using smoothing spline models. The implementation of the smoothing spline model was done in R software using *smooth*.*spline* function with four knots. The models were then used to extrapolate the daily values of *R*_*t*_ outside of Jakarta based on the province-level 7-day rolling average of changes in non-residential mobility (Google Community Mobility Reports^5^).

### S4. Estimating movement matrices from CDRs data

District level (city and municipality, 115 in total) movement matrices (*M*) prior to the pandemic for the normal (or non-Ramadan) (*M*_*N*_) and Ramadan (*M*_*R*_) periods were calculated from CDRs data. Each element of the matrix (*m*_*i, j*_) represents the proportion of days spent by residents of district *i* in district *j* over the year.

The daily position of each user is described as the district where the most frequent mobile phone usage happened over the period of a single day. All users were assumed to be active from the first day to the last day of their phone usage. On days where the user was not active (no phone activities recorded), the position of the user was assumed to be the same as the previous day. The primary residence of each user is defined as the district where the users spent most of their days over their ‘active’ period. Based on their daily locations and primary residences, we then calculated the proportion of days spent of people from district *i* in district *j. M*_*R*_ was estimated using data of August and September 2011 (period of the Ramadan month, Eid celebration, and national holidays period). *M*_*N*_ was estimated using the rest of the data that were not used to estimate *M*_*R*_ (May 2011 - July 2011 and October 2011 - April 2012 periods).

All districts in the Java island were represented as a single row in each matrix with exceptions for districts in Jakarta province which were represented as an aggregated single (*i* = 1). Outside Java movements were represented as a single row (*i* = 2). **Table S1** shows a complete list of districts (including Jakarta and outside Java) and each respective index in the movement matrix.

### S5. Metapopulation Model

#### Metapopulation model of COVID-19 spread in Java

We developed a metapopulation model to simulate the spread of COVID-19 in Java. Each patch in the metapopulation model represents districts (*i* = 1, 2, …, 115) listed in **Table S1**. For each patch, stochastic differential equations representing a Susceptible-Exposed-Infected-Recovered (SEIR) model were implemented (overall structure in **Figure S3**). The equations are as follows:

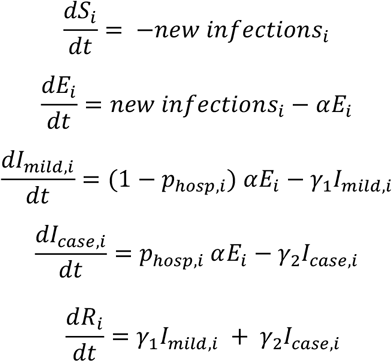

where *new infections*_*i*_ is the number of new infections in each patch based on the stochastic adaptation of the metapopulation transmission model by Keeling et al.^12^, *α* is the per-capita transition rate reflecting the mean duration of latent period, *p*_*hosp*_ is the probability of having severe illness and needing hospitalisations, *p*_*critical*|*hosp*_ is the probability of needing critical care if hospitalised, and *γ*_1_ and *γ*_2_ are the per-capita transition rate reflecting the mean duration of infectiousness of mild and severe infections, respectively. The full model parameter descriptions and specifications are available in **Table S2**. To calculate *new infections*_*i*_, we firstly need to calculate the district-level force of infections *λ*_*i*_ that accounts for inter-district movements of both susceptible individuals (that might acquire infections in other districts) and infected individuals (that might infect people in other districts) based on the inter-patch connectivities (the movement matrix, *M*, accounting daily changes in mobility - see **section S4**). The total number of infected individuals that are contributing to infections in district *i, I*_*tot, i*_, is calculated by:

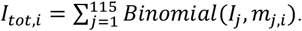

The transmission rate for each district is calculated by:

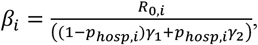

where *R*_0,*i*_ is the value of *R*_*t*_ estimated for the period of maximum mobility recorded within Google Mobility data.

Hence,:

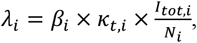

where *N*_*i*_ is the total population of each district/patch and k_*t, i*_ is the daily ratio between the estimated *R*_*t, i*_ values based on the spline model estimates and the respective *R*_0,*i*_, representing the relative changes in the daily transmission rate. We assumed no transmission contributed to and from outside Java but we still allow movement to and from that patch (*i* = 2) which implies both *I*_2_ and *λ*_2_ are always 0.

*new infections*_*i*_ are then calculated as:

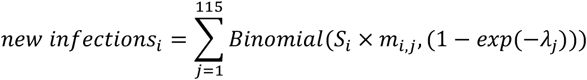

For each severe infection needing hospitalisation (*I*_*case*_), the case was either deemed a critical case (i.e., indicated to require an ICU bed) with probability *p*_*critical*|*hosp*_ and otherwise, non-critical (i.e., indicated to require an isolation bed) with associated probability of death (*p*_*death*|*non critical*_ and *p*_*death*|*critical*_, diagram in **Figure S3B**). *p*_*hosp*_, *p*_*critical*|*hosp*_, *p*_*death*|*non critical*_ and *p*_*death*|*critical*_ were obtained for each district as the average values estimated within simulations from the squire package^13^, taking age-specific demography using district-level census data and, in the absence of equivalent data from Java, mixing patterns based upon a contact survey from Shanghai province, China as an example of contact patterns within a UMIC Asian country and province containing a megacity.

The model was simulated with 100 replicates by seeding initial cases in Jakarta, and Kota Surabaya (East Java’s capital) – both have international airports receiving travellers from China – on 7^th^ January 2020 (arbitrarily selected) with 100 replicates. Initial cases in Jakarta were set to 12, obtained through calibration to provide simulated deaths trends bounded by the interval between reported deaths and C19P funerals. Initial cases in Surabaya were drawn according to a binomial draw assuming an underlying importation rate of 60% of that in Jakarta^14^, random numbers based on the binomial distribution were sampled for each replicate.

We assumed different transmission scenarios of districts classified as rural and urban districts. We classified an urban/rural status to each district based upon the urban/rural classification of the majority of villages within the district. We assigned *R*_*t, i*_ values for all districts to be the province-level value of *R*_*t, i*_ in which each district is located. We then explore the possibilities of rural districts to have different *R*_*t, i*_ levels, ranging from 100% to 60% of province-level *R*_*t, i*_. In the results section of the main text, we show simulation results assuming *R*_*t, i*_ in rural districts to be 90% of *R*_*t, i*_ in urban districts. We also ran the model for several different counterfactual scenarios. The list of assumed transmission scenarios and the counterfactual scenarios were shown in **Table S3 & S4**.

During the period of Ramadan and Eid festivals, 24^th^ April 2020 up to 7^th^ June 2020, the Ramadan movement matrix (*M*_*R*_) was used as the baseline movement matrix. In the other period, the non-Ramadan movement matrix (*M*_*N*_) was used as the baseline movement matrix. As a baseline assumption, throughout the simulation, the proportions of people spending their time in other districts (*m*_, *j*_ where *i* ≠ *j*) were adjusted by the province-level changes in mobility over time, with reductions in larger-scale movement outside the province assumed to be higher than those within the province according to an odds ratio (OR) of 2 within our default scenario.

For each scenario, we also devised a metric to assess the extent to which the epidemic would be likely to strain available healthcare resources over time given patterns of spatial spread and disparities in healthcare supply by district. This metric was defined by determining the number of available beds for each individual newly requiring hospitalisation each day within the model by subtracting the number of hospital beds required in the simulation from the total hospital beds capacity available at the district-level obtained from the Online Healthcare Facilities website.^6^

**Figure S3.**
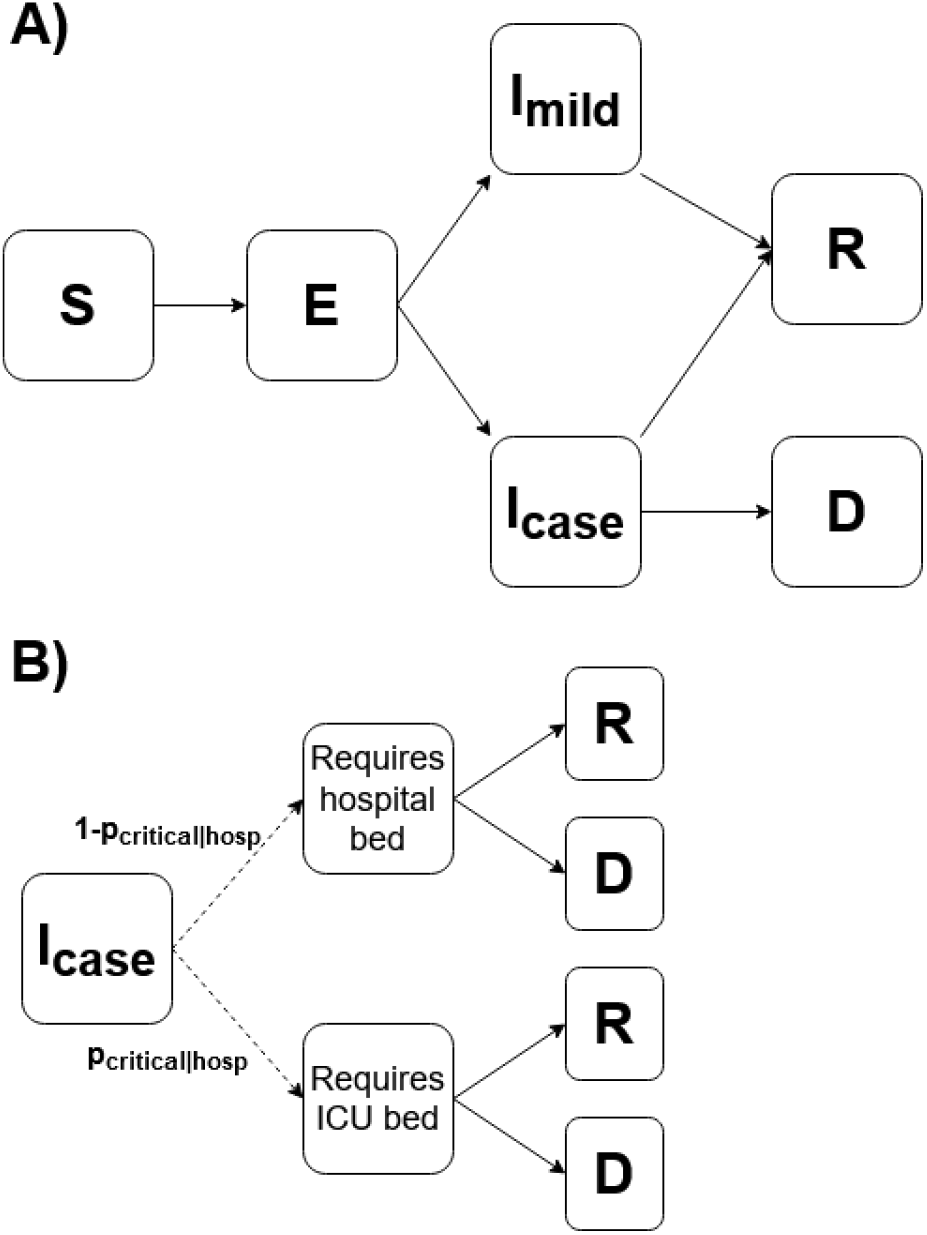
SEIR model structure. **A)** The SEIR structure for each patch in the metapopulation model. Susceptible individuals (***S***), if infected, progress to the exposed compartment (***E***), having their latent period of infections. Then, those individuals will either develop mild symptoms (***I***_***mild***_) or severe symptoms requiring hospitalisations (*I*_*case*_). Those who developed severe symptoms may have two possible outcomes: recovery (*R*) or death (*D*); **B)** The pathway of infections requiring hospitalisations (*I*_*case*_). Each infection with severe symptoms may only need a standard hospital bed or may develop worse conditions that require critical care (ICU bed). Each of those cases treated in both critical and non-critical care may recover or die based on specific probabilities. Dashed lines denote probabilistic pathways, not rates of transition.

**Table S1.** List of districts, districts’ indexes, and probability of disease severity and outcomes for the metapopulation model.

| Index no. | District | Province | $p_{hosp,i}$ | $p_{critical\vert hosp,i}$ | $p_{death\vert non\ critical,i}$ |
| --- | --- | --- | --- | --- | --- |
| 1 | Jakarta | Jakarta | 0.029 | 0.185 | 0.054 |
| 2 | Outside Java | Outside Java | 0.027 | 0.195 | 0.058 |
| 3 | Bogor | West Java | 0.026 | 0.190 | 0.056 |
| 4 | Sukabumi | West Java | 0.031 | 0.216 | 0.065 |
| 5 | Cianjur | West Java | 0.031 | 0.212 | 0.064 |
| 6 | Bandung | West Java | 0.028 | 0.201 | 0.060 |
| 7 | Garut | West Java | 0.030 | 0.219 | 0.067 |
| 8 | Tasikmalaya | West Java | 0.034 | 0.228 | 0.070 |
| 9 | Ciamis | West Java | 0.039 | 0.234 | 0.072 |
| 10 | Kuningan | West Java | 0.036 | 0.233 | 0.072 |
| 11 | Cirebon | West Java | 0.030 | 0.209 | 0.063 |
| 12 | Majalengka | West Java | 0.036 | 0.229 | 0.070 |
| 13 | Sumedang | West Java | 0.036 | 0.234 | 0.073 |
| 14 | Indramayu | West Java | 0.033 | 0.210 | 0.063 |
| 15 | Subang | West Java | 0.035 | 0.224 | 0.069 |
| 16 | Purwakarta | West Java | 0.029 | 0.206 | 0.061 |
| 17 | Karawang | West Java | 0.030 | 0.200 | 0.059 |
| 18 | Bekasi | West Java | 0.025 | 0.177 | 0.051 |
| 19 | Bandung Barat | West Java | 0.030 | 0.214 | 0.065 |
| 20 | Pangandaran | West Java | 0.038 | 0.231 | 0.071 |
| 21 | Kota Bogor | West Java | 0.029 | 0.195 | 0.058 |
| 22 | Kota Sukabumi | West Java | 0.031 | 0.212 | 0.064 |
| 23 | Kota Bandung | West Java | 0.030 | 0.199 | 0.059 |
| 24 | Kota Cirebon | West Java | 0.031 | 0.201 | 0.060 |
| 25 | Kota Bekasi | West Java | 0.026 | 0.165 | 0.047 |
| 26 | Kota Depok | West Java | 0.027 | 0.176 | 0.051 |
| 27 | Kota Cimahi | West Java | 0.028 | 0.191 | 0.056 |
| 28 | Kota Tasikmalaya | West Java | 0.031 | 0.207 | 0.062 |
| 29 | Kota Banjar | West Java | 0.036 | 0.227 | 0.070 |
| 30 | Cilacap | Central Java | 0.035 | 0.232 | 0.072 |
| 31 | Banyumas | Central Java | 0.036 | 0.236 | 0.073 |
| 32 | Purbalingga | Central Java | 0.035 | 0.233 | 0.072 |
| 33 | Banjarnegara | Central Java | 0.035 | 0.229 | 0.071 |
| 34 | Kebumen | Central Java | 0.037 | 0.247 | 0.078 |
| 35 | Purworejo | Central Java | 0.040 | 0.254 | 0.081 |
| 36 | Wonosobo | Central Java | 0.035 | 0.230 | 0.071 |
| 37 | Magelang | Central Java | 0.036 | 0.236 | 0.073 |
| 38 | Boyolali | Central Java | 0.038 | 0.248 | 0.078 |
| 39 | Klaten | Central Java | 0.039 | 0.249 | 0.079 |
| 40 | Sukoharjo | Central Java | 0.035 | 0.233 | 0.072 |
| 41 | Wonogiri | Central Java | 0.044 | 0.260 | 0.084 |
| 42 | Karanganyar | Central Java | 0.036 | 0.235 | 0.073 |
| 43 | Sragen | Central Java | 0.038 | 0.244 | 0.077 |
| 44 | Grobogan | Central Java | 0.035 | 0.230 | 0.071 |
| 45 | Blora | Central Java | 0.037 | 0.240 | 0.075 |
| 46 | Rembang | Central Java | 0.034 | 0.225 | 0.069 |
| 47 | Pati | Central Java | 0.036 | 0.233 | 0.072 |
| 48 | Kudus | Central Java | 0.031 | 0.206 | 0.062 |
| 49 | Jepara | Central Java | 0.032 | 0.219 | 0.067 |
| 50 | Demak | Central Java | 0.030 | 0.210 | 0.063 |
| 51 | Semarang | Central Java | 0.035 | 0.234 | 0.073 |
| 52 | Temanggung | Central Java | 0.036 | 0.231 | 0.071 |
| 53 | Kendal | Central Java | 0.033 | 0.221 | 0.067 |
| 54 | Batang | Central Java | 0.033 | 0.217 | 0.066 |
| 55 | Pekalongan | Central Java | 0.031 | 0.217 | 0.066 |
| 56 | Pemalang | Central Java | 0.033 | 0.224 | 0.068 |
| 57 | Tegal | Central Java | 0.032 | 0.221 | 0.068 |
| 58 | Brebes | Central Java | 0.032 | 0.222 | 0.068 |
| 59 | Kota Magelang | Central Java | 0.036 | 0.229 | 0.071 |
| 60 | Kota Surakarta | Central Java | 0.033 | 0.220 | 0.067 |
| 61 | Kota Salatiga | Central Java | 0.033 | 0.227 | 0.070 |
| 62 | Kota Semarang | Central Java | 0.030 | 0.205 | 0.061 |
| 63 | Kota Pekalongan | Central Java | 0.030 | 0.202 | 0.060 |
| 64 | Kota Tegal | Central Java | 0.031 | 0.210 | 0.063 |
| 65 | Kulon Progo | Yogyakarta | 0.040 | 0.248 | 0.078 |
| 66 | Bantul | Yogyakarta | 0.035 | 0.232 | 0.072 |
| 67 | Gunung Kidul | Yogyakarta | 0.042 | 0.253 | 0.080 |
| 68 | Sleman | Yogyakarta | 0.033 | 0.223 | 0.068 |
| 69 | Kota Yogyakarta | Yogyakarta | 0.032 | 0.215 | 0.065 |
| 70 | Pacitan | East Java | 0.042 | 0.252 | 0.080 |
| 71 | Ponorogo | East Java | 0.041 | 0.247 | 0.078 |
| 72 | Trenggalek | East Java | 0.039 | 0.240 | 0.075 |
| 73 | Tulungagung | East Java | 0.038 | 0.237 | 0.074 |
| 74 | Blitar | East Java | 0.039 | 0.243 | 0.076 |
| 75 | Kediri | East Java | 0.036 | 0.231 | 0.071 |
| 76 | Malang | East Java | 0.036 | 0.228 | 0.070 |
| 77 | Lumajang | East Java | 0.036 | 0.222 | 0.068 |
| 78 | Jember | East Java | 0.035 | 0.224 | 0.068 |
| 79 | Banyuwangi | East Java | 0.037 | 0.230 | 0.071 |
| 80 | Bondowoso | East Java | 0.038 | 0.227 | 0.070 |
| 81 | Situbondo | East Java | 0.036 | 0.218 | 0.067 |
| 82 | Probolinggo | East Java | 0.034 | 0.217 | 0.066 |
| 83 | Pasuruan | East Java | 0.031 | 0.201 | 0.060 |
| 84 | Sidoarjo | East Java | 0.029 | 0.190 | 0.056 |
| 85 | Mojokerto | East Java | 0.033 | 0.211 | 0.063 |
| 86 | Jombang | East Java | 0.034 | 0.223 | 0.069 |
| 87 | Nganjuk | East Java | 0.037 | 0.231 | 0.072 |
| 88 | Madiun | East Java | 0.041 | 0.241 | 0.075 |
| 89 | Magetan | East Java | 0.043 | 0.251 | 0.080 |
| 90 | Ngawi | East Java | 0.040 | 0.236 | 0.074 |
| 91 | Bojonegoro | East Java | 0.037 | 0.229 | 0.071 |
| 92 | Tuban | East Java | 0.035 | 0.223 | 0.068 |
| 93 | Lamongan | East Java | 0.037 | 0.226 | 0.069 |
| 94 | Gresik | East Java | 0.031 | 0.201 | 0.059 |
| 95 | Bangkalan | East Java | 0.032 | 0.227 | 0.069 |
| 96 | Sampang | East Java | 0.029 | 0.211 | 0.064 |
| 97 | Pamekasan | East Java | 0.031 | 0.211 | 0.064 |
| 98 | Sumenep | East Java | 0.036 | 0.218 | 0.066 |
| 99 | Kota Kediri | East Java | 0.033 | 0.212 | 0.064 |
| 100 | Kota Blitar | East Java | 0.035 | 0.225 | 0.069 |
| 101 | Kota Malang | East Java | 0.031 | 0.211 | 0.064 |
| 102 | Kota Probolinggo | East Java | 0.032 | 0.207 | 0.062 |
| 103 | Kota Pasuruan | East Java | 0.030 | 0.203 | 0.060 |
| 104 | Kota Mojokerto | East Java | 0.033 | 0.209 | 0.063 |
| 105 | Kota Madiun | East Java | 0.037 | 0.227 | 0.070 |
| 106 | Kota Surabaya | East Java | 0.030 | 0.194 | 0.057 |
| 107 | Kota Batu | East Java | 0.034 | 0.221 | 0.068 |
| 108 | Pandeglang | Banten | 0.029 | 0.205 | 0.061 |
| 109 | Lebak | Banten | 0.028 | 0.194 | 0.057 |
| 110 | Tangerang | Banten | 0.025 | 0.172 | 0.049 |
| 111 | Serang | Banten | 0.027 | 0.185 | 0.054 |
| 112 | Kota Tangerang | Banten | 0.026 | 0.161 | 0.046 |
| 113 | Kota Cilegon | Banten | 0.026 | 0.167 | 0.048 |
| 114 | Kota Serang | Banten | 0.025 | 0.167 | 0.048 |
| 115 | Kota Tangerang Selatan | Banten | 0.027 | 0.169 | 0.048 |

**Table S2.** Model parameters descriptions and values for the metapopulation model.

| Parameter | Symbol | Value | Description |
| --- | --- | --- | --- |
| Transmission rate | $\beta_i$ | - | Calculated from $R_{0,i}$. |
| Basic reproduction number | $R_{0,i}$ | - | Estimated from the maximum values of the smoothing spline models between mobility changes and $R_{t,funerals}$. |
| Relative changes in daily transmission rate | $\kappa_i$ | - | The ratio between $R_{0,i}$ and daily estimated $R_{t,i}$ based on spline smoothing models. |
| Mean latent period | $1/\alpha$ | 4.6 days | Estimated as 5.1 days<sup>15</sup> with 0.5 days accounted as a pre-symptomatic period of infectiousness. |
| Mean duration of infectiousness of mild infections | $1/\gamma_1$ | 2.1 days | 0.5 days infectiousness period prior to symptoms included which in combination with mean duration of severe illness gives a mean serial interval of 6.75 days.<sup>11</sup> |
| Mean duration of infectiousness of severe infections | $1/\gamma_2$ | 4.5 days | Mean onset to admission to hospital of 4 days, as used in squire model<sup>16</sup>, based on unpublished analysis of data from the ICNARC study<sup>17</sup> and includes 0.4 days of infectiousness prior to symptoms. |
| Probability of having severe illness, needing hospitalisations | $p_{hosp,i}$ | **Table S1** | For each district, the probability was calculated by running a full unmitigated epidemic in squire package<sup>13</sup>. The proportion of people needing hospitalisations were calculated based on the simulations. |
| Probability of needing critical care, if hospitalised | $p_{critical\vert hosp,i}$ | **Table S1** | For each district, the probability was calculated by running a full unmitigated epidemic in squire package<sup>13</sup>. The proportion of people needing hospitalisations were calculated based on the simulations. |
| Probability of death of severe illness that does not need critical care | $p_{death\vert non\ critical,i}$ | **Table S1** | For each district, the probability was calculated by running a full unmitigated epidemic in squire package.<sup>13</sup> The proportion of people needing hospitalisations were calculated based on the simulations. |
| Probability of death of severe illness that needs critical care | $p_{death\vert critical,i}$ | 0.5 | Probability of death from severe illness needing critical care based on the data of the ICNARC study in the UK<sup>17</sup>. |

**Table S3.** List of assumed transmission scenarios for metapopulation model simulations.

| Transmission scenarios |  |  |
| --- | --- | --- |
| Scenario name | Details | Results shown in |
| Rural 100% | $R_0$ and $R_t$ of urban and rural districts in each province were assumed to be the same. | **Figure S6** |
| Rural 90% | $R_0$ and $R_t$ of rural districts were assumed to be 90% of the province level $R_0$ and $R_t$. Used as the main transmission scenario shown in the main text. | **Figure 4; Figure S6** |
| Rural 75% | $R_0$ and $R_t$ of rural districts were assumed to be 75% of the province level $R_0$ and $R_t$. | **Figure S6** |
| Rural 60% | $R_0$ and $R_t$ of rural districts were assumed to be 60% of the province level $R_0$ and $R_t$. | **Figure S6** |

**Table S4.**
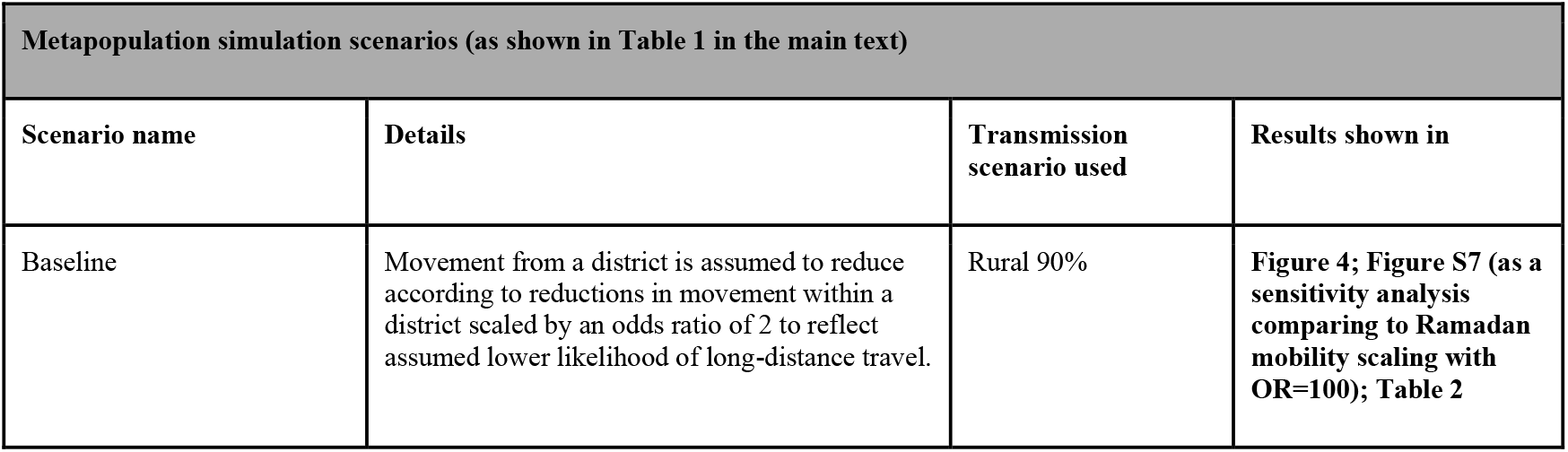
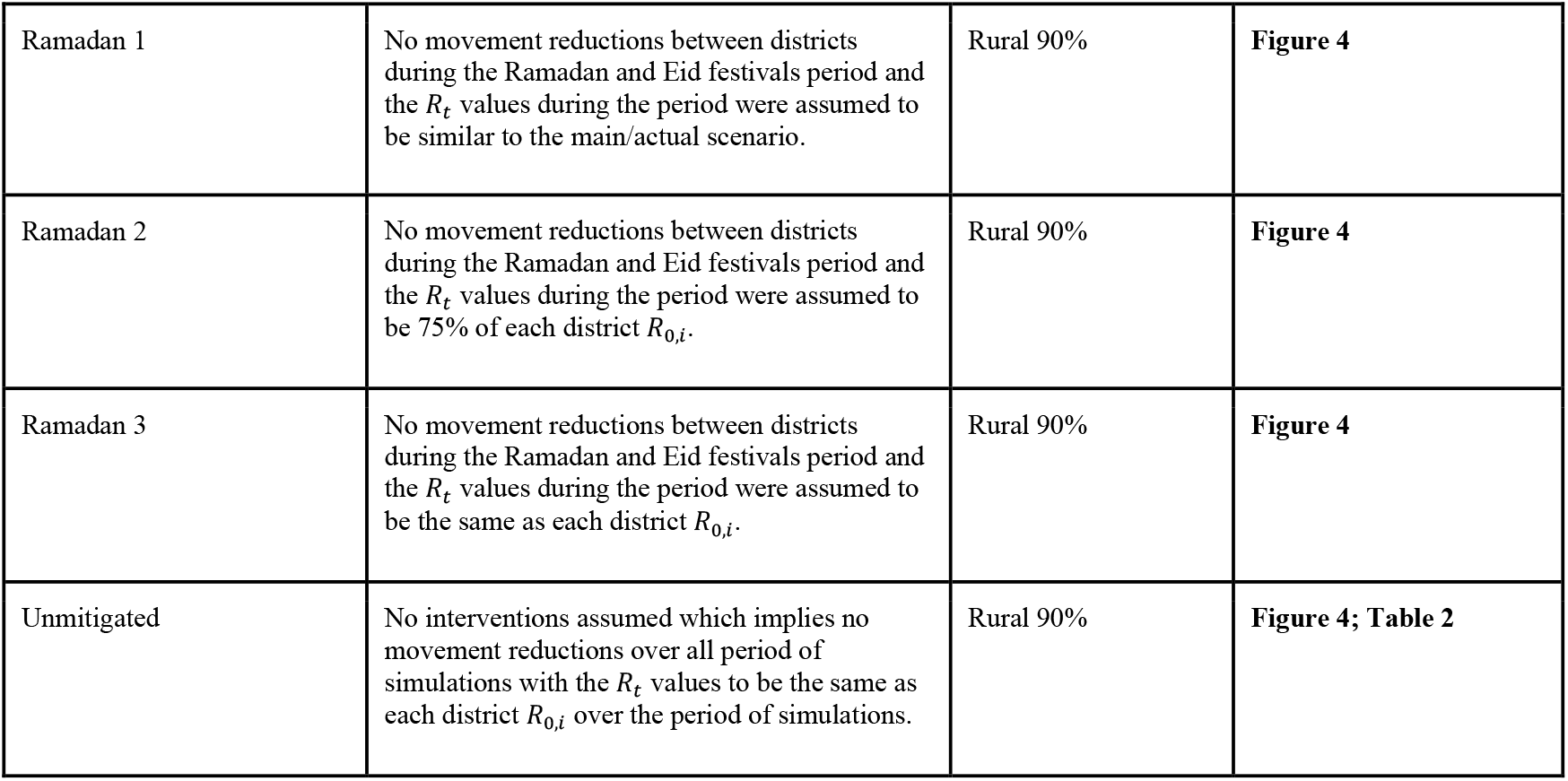
List of counterfactual scenarios for metapopulation model simulations.

| Metapopulation simulation scenarios (as shown in Table 1 in the main text) |  |  |  |
| --- | --- | --- | --- |
| Scenario name | Details | Transmission scenario used | Results shown in |
| Baseline | Movement from a district is assumed to reduce according to reductions in movement within a district scaled by an odds ratio of 2 to reflect assumed lower likelihood of long-distance travel. | Rural 90% | **Figure 4; Figure S7 (as a sensitivity analysis comparing to Ramadan mobility scaling with OR=100); Table 2** |
| Ramadan 1 | No movement reductions between districts during the Ramadan and Eid festivals period and the $R_t$ values during the period were assumed to be similar to the main/actual scenario. | Rural 90% | Figure 4 |
| Ramadan 2 | No movement reductions between districts during the Ramadan and Eid festivals period and the $R_t$ values during the period were assumed to be 75% of each district $R_{0,i}$. | Rural 90% | Figure 4 |
| Ramadan 3 | No movement reductions between districts during the Ramadan and Eid festivals period and the $R_t$ values during the period were assumed to be the same as each district $R_{0,i}$. | Rural 90% | Figure 4 |
| Unmitigated | No interventions assumed which implies no movement reductions over all period of simulations with the $R_t$ values to be the same as each district $R_{0,i}$ over the period of simulations. | Rural 90% | Figure 4; Table 2 |

### S6. Model fitting to confirmed and suspected COVID-19 deaths and future projection scenarios in all provinces in Java

#### Estimating the number of deaths from suspected and probable cases in Java provinces

Jakarta reported a time series dataset of the province daily C19P funerals in their official COVID-19 tracker website (https://corona.jakarta.go.id/id/data-pemantauan)^1^ which includes confirmed/reported and probable COVID-19 deaths (both combined were then defined as suspected deaths). Whilst for the other five provinces in Java, daily time series data of probable deaths are not available. WHO Indonesia situation reports provide a weekly summary of confirmed and probable deaths (which both combined become suspected deaths) in all provinces in Java since the end of May 2020 (**Figure S8**)^4^ We collated these data and calculated the proportion of confirmed deaths from suspected COVID-19 deaths for each province (*ρ*_*i*_ with *i* as each province index).

For all days from 1^st^ March 2020, up to 7^th^ December 2020, we simulated the number of probable deaths in five Java provinces other than Jakarta. Firstly, we aggregated the daily confirmed deaths in each province to a weekly period (*D*_*i, t*_ with *t* as the weekly time window). For each weekly time window *t*, using Negative Binomial distribution, we simulated the number of probable deaths (*O*_*i, t*_) in each province 10 times:

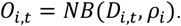

For each simulated *O*_*i, t*_, we simulated the spread of the weekly total estimated probable deaths into daily estimated probable deaths using Multinomial distribution, assuming equal probability for all days during the week 10 times:

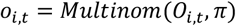

where *π* is a vector of length 7, where each value is 1/7.

The simulations resulted in 100 samples of estimated daily probable deaths in each province. Adding the simulated daily probable deaths to the daily confirmed deaths, we obtained 100 samples of the estimated number of daily suspected COVID-19 deaths in five provinces in Java other than Jakarta.

The daily suspected COVID-19 deaths are then defined, for Jakarta, as the daily C19P funerals, and for five other Java provinces as the median of the estimated number of the daily suspected COVID-19 deaths.

#### Model fitting

Using the framework developed in the Imperial College COVID-19 LMIC reports^18^, we fit the model to both the daily COVID-19 confirmed deaths and the daily COVID-19 suspected deaths data for each province in Java, estimating both a province-specific *R*_0_ and epidemic start date. To provide model fits that are agnostic to the mobility profiles in each province, we model the time-varying reproduction number, *R*_*t*_, using a series of pseudo-random walk parameters, which alter the transmission every 2-weeks, with *R*_*t*_ given by:

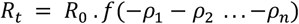

Where *f*(*x*) = 2 *exp*(*x*)/ (1 + *exp*(*x*)), i.e., twice the inverse logit function, which has been used in previous models to capture the impact of mobility data on transmission^19^. Each *ρ* parameter is introduced two weeks after the previous parameter, serving to capture changes in transmission over time. More specifically, each *ρ* parameter is set equal to 0 for each day prior to its start date. For example, *ρ*_1_ is the change in transmission, which is set to start at the beginning of the epidemic. The estimated value for *ρ*_1_ is then maintained for all future time points. *ρ*_2_ is the second change in transmission, which starts 14 days after the epidemic start date, i.e., is equal to 0 prior to this. The last mobility independent change in transmission, *ρ*_*n*_ is maintained for the last 4 weeks prior to the current day to reflect our inability to estimate the effect size of this parameter due to the approximate 21 day delay between infection and death.^16^

The model fitting results were presented in **Figure S9 & S10**. Based on the fitted models in all provinces considering different types of deaths data, we estimated the attack rate at the province level and Java level.

#### Future projection scenarios

Using the fitted models, some future scenarios were explored based on the assumed values of the reproduction number under control, *R*_*c*_, defined similarly to *R*_0_ as the average number of secondary infections within an entirely susceptible population but incorporating the impact of NPIs (and, equivalently, *R*_*t*_ but not incorporating the effects of population-level immunity such that *R*_0_ > *R*_*c*_ > *R*_*t*_). Moreover, as with *R*_0_, *R*_*c*_>1 can lead to *R*_*t*_ < 1 and a declining epidemic if there exist sufficient levels of naturally acquired immunity within the population. We simulated forward projections based on scenarios described in **Table S5**.

**Table S5.**
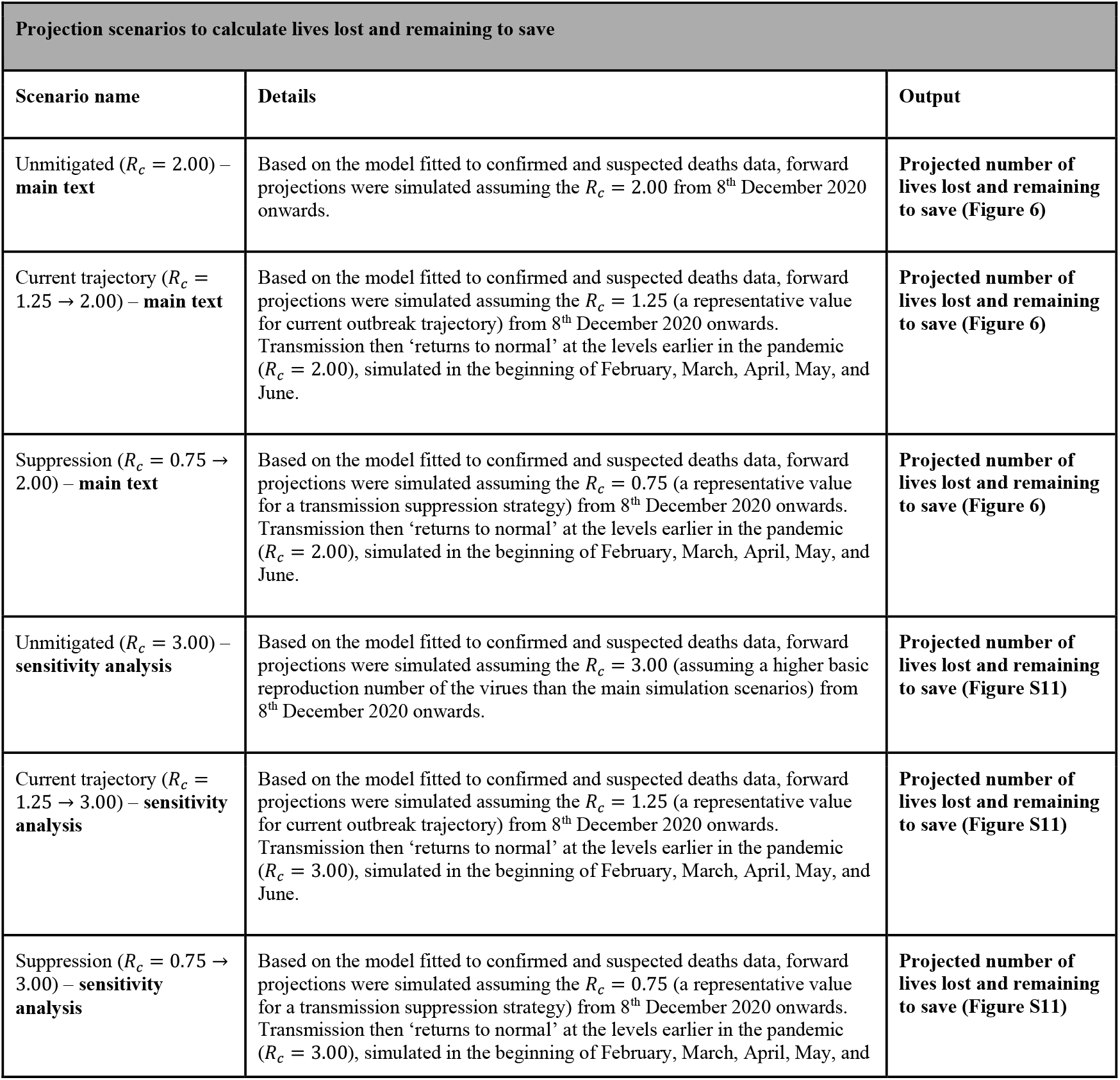
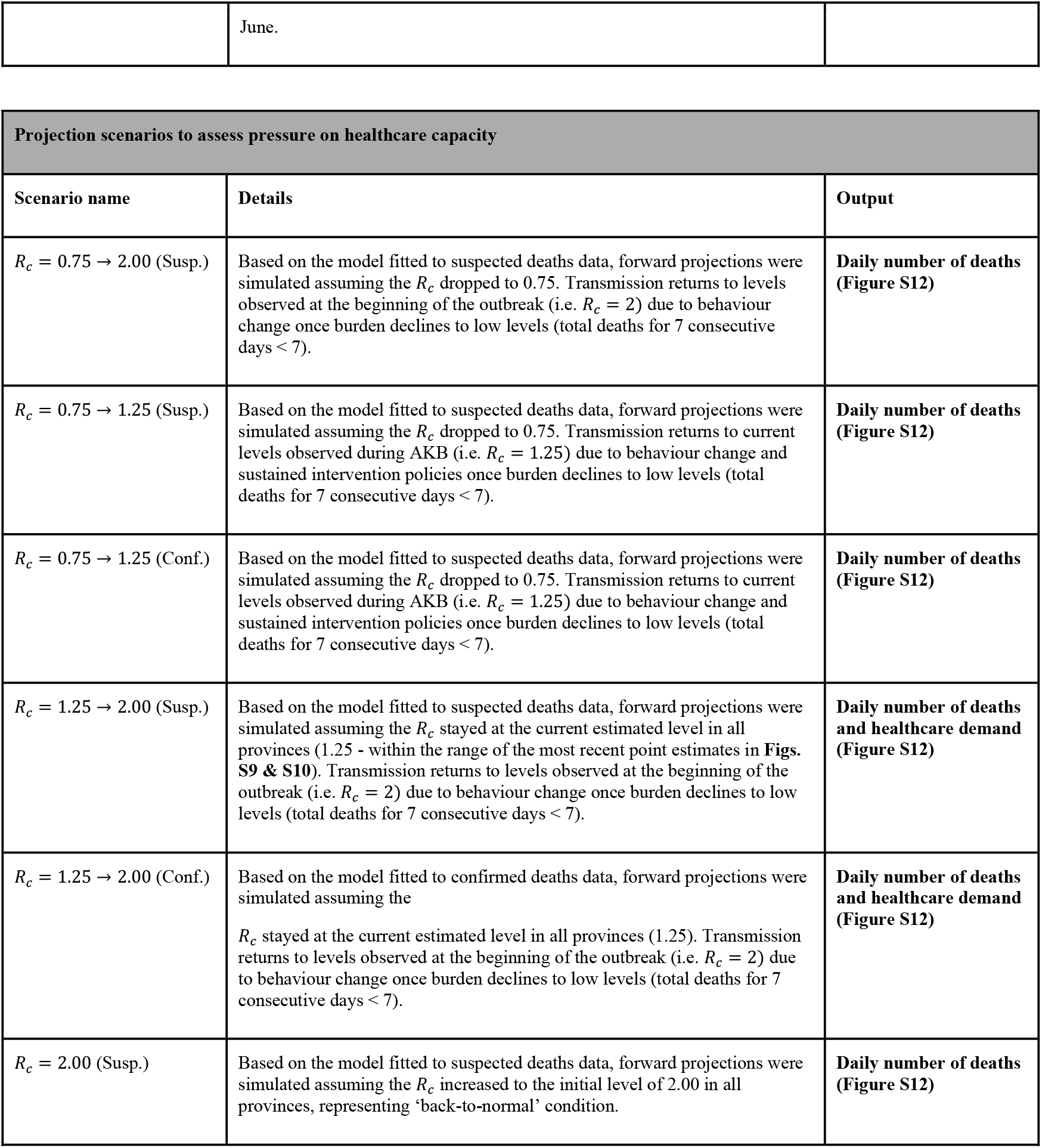
List of future projection scenarios.

**Figure S4.**
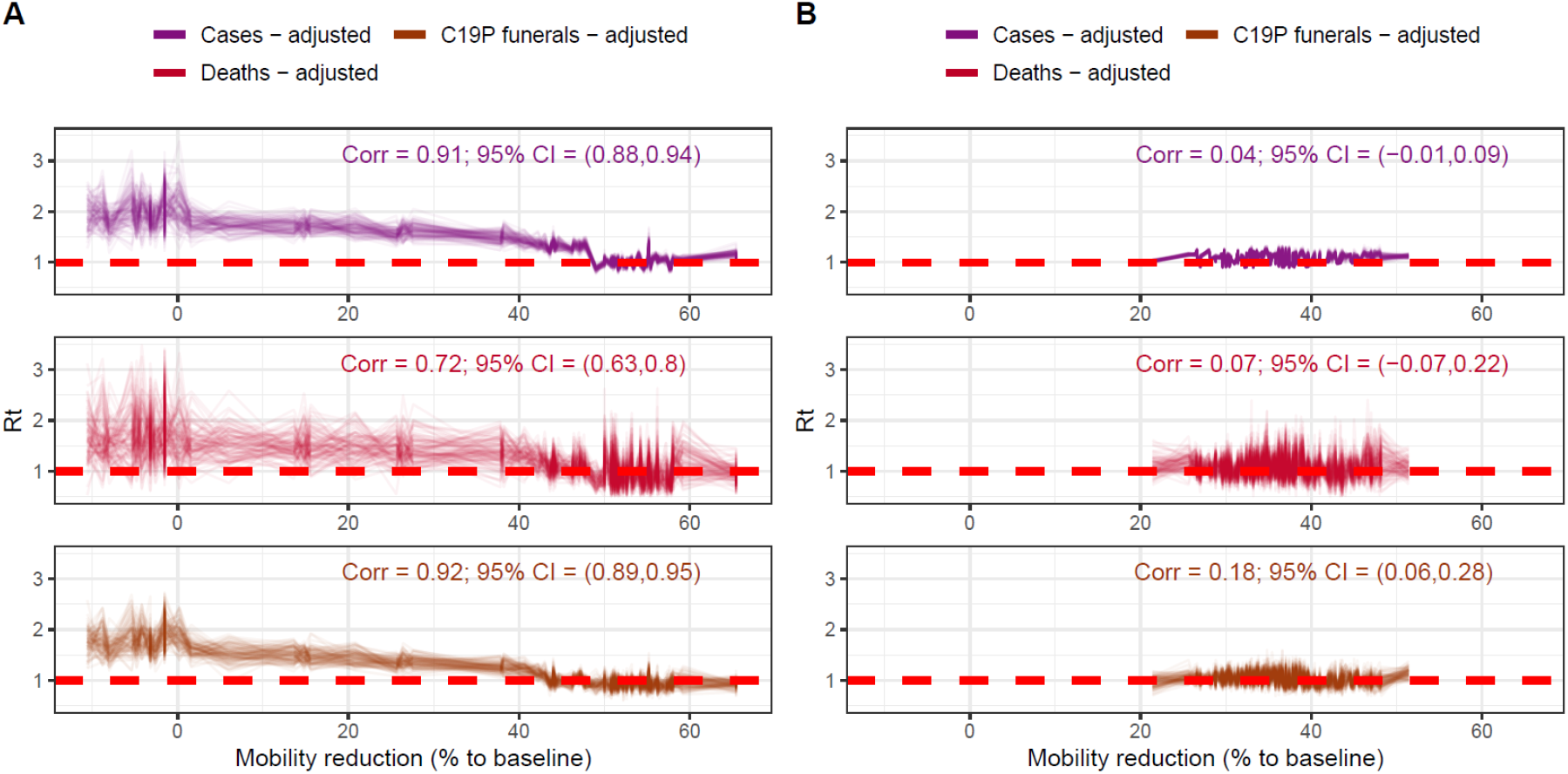
Decorrelations between estimated *R*_*t, funerals*_ in Jakarta and mobility changes based on *R*_*t, funerals*_ estimates before AKB (the ‘new normal’) **(A)** and after AKB **(B)**.

**Figure S5.**
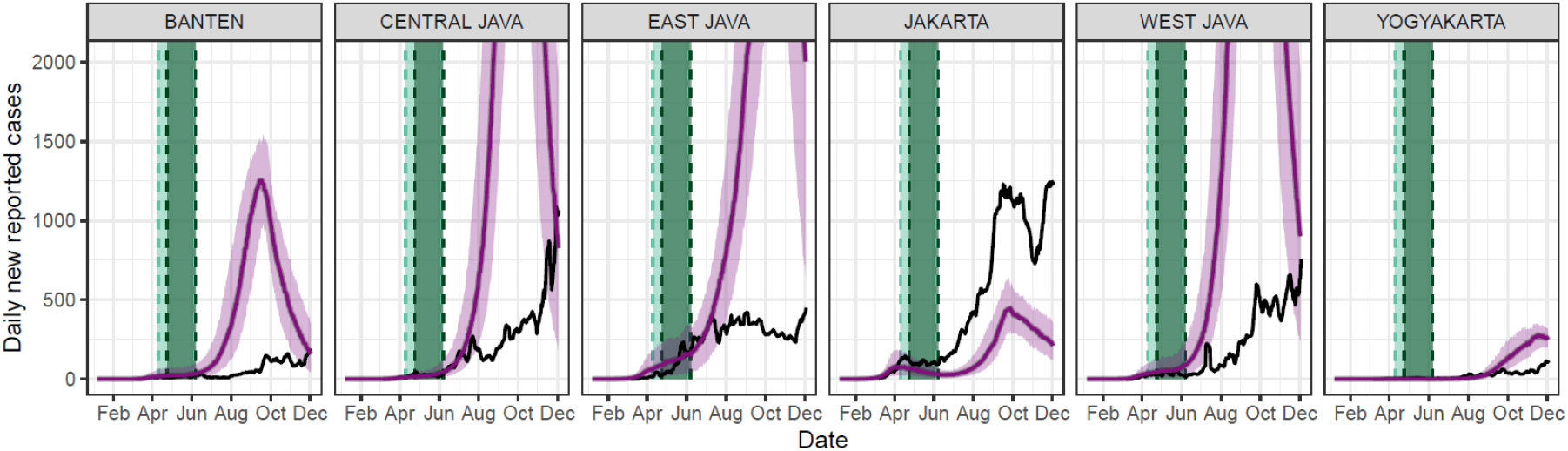
Comparison of model simulations and observed daily new reported cases from COVID-19. Coloured lines and their shaded areas denote model simulation outputs with their respective uncertainties (95% level) while black lines denote observed data.

**Figure S6.**
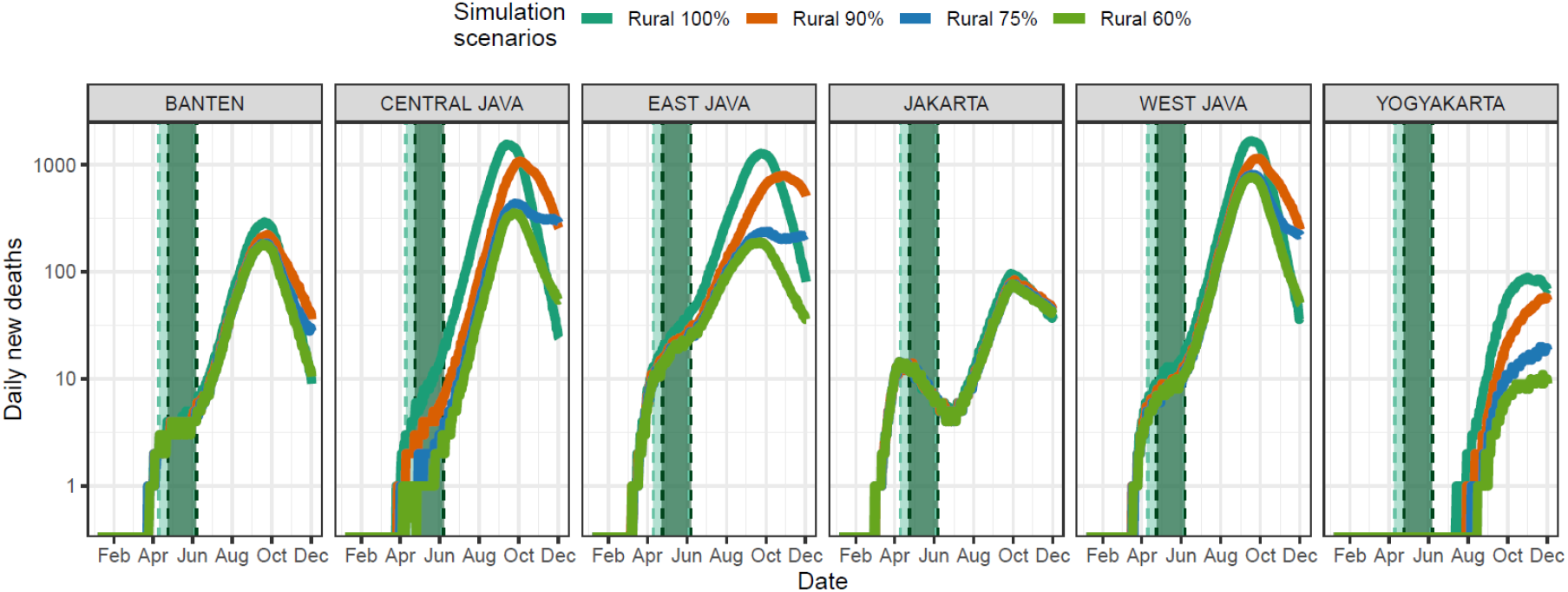
Simulated daily new deaths comparing rural transmission scenarios at the province-level.

**Figure S7.**
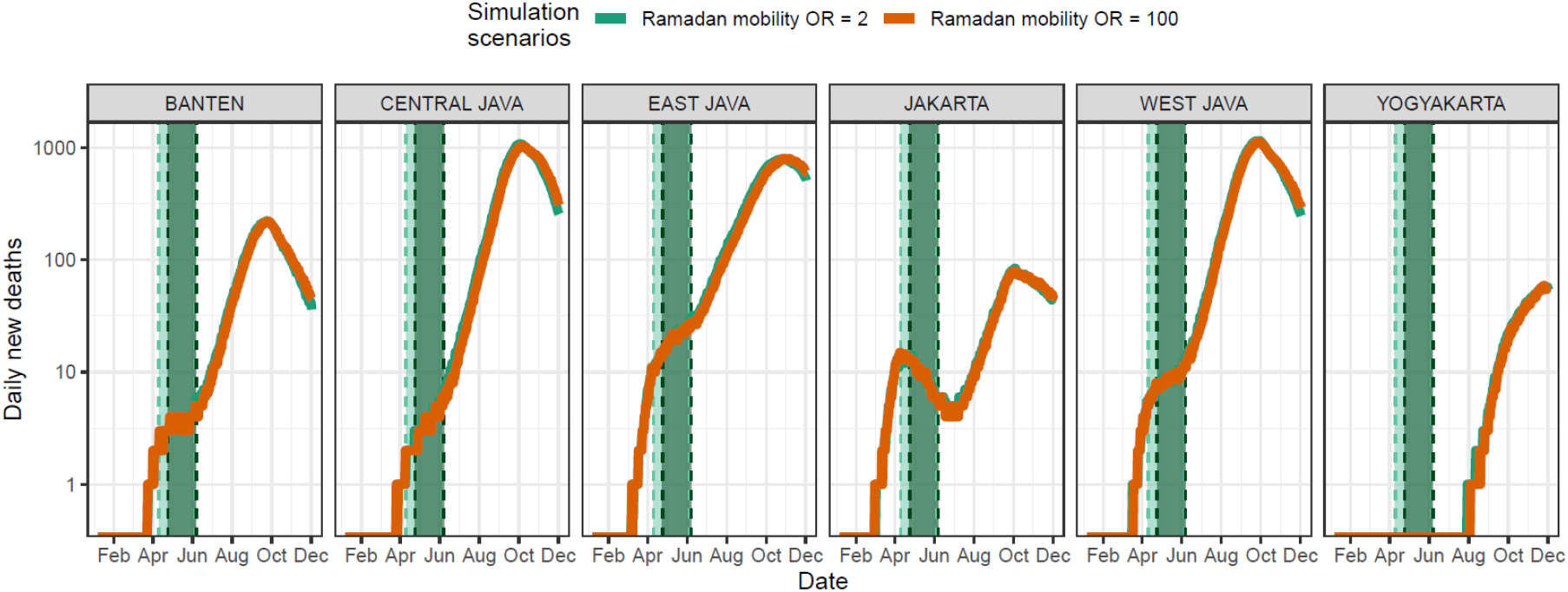
Sensitivity analysis of assumed impact of between-district movement restrictions at the province-level.

**Figure S8.**
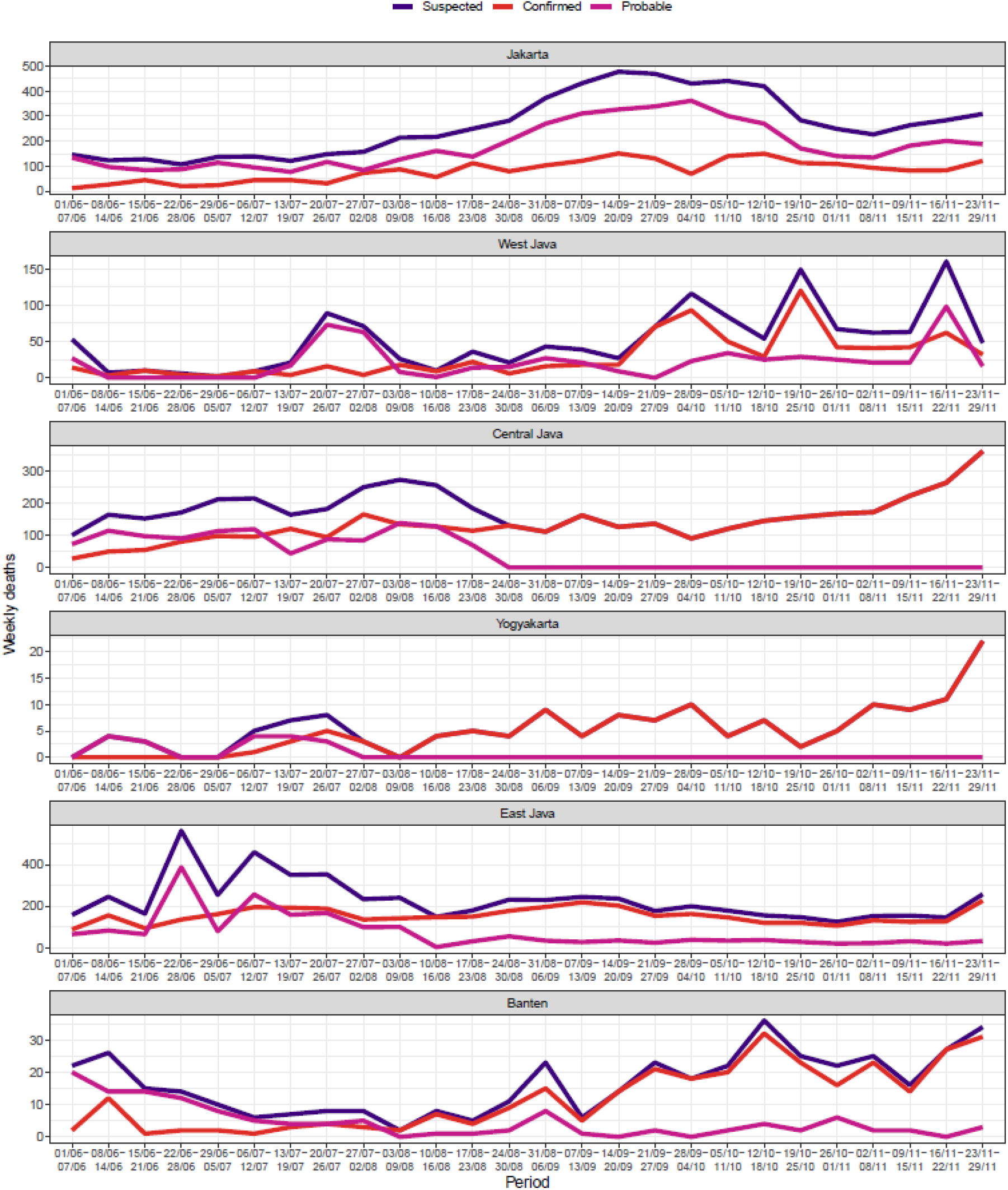
Weekly aggregated reported/confirmed and probable deaths in Java collated from WHO COVID-19 Indonesia situation reports 13-36.^4^

**Figure S9.**
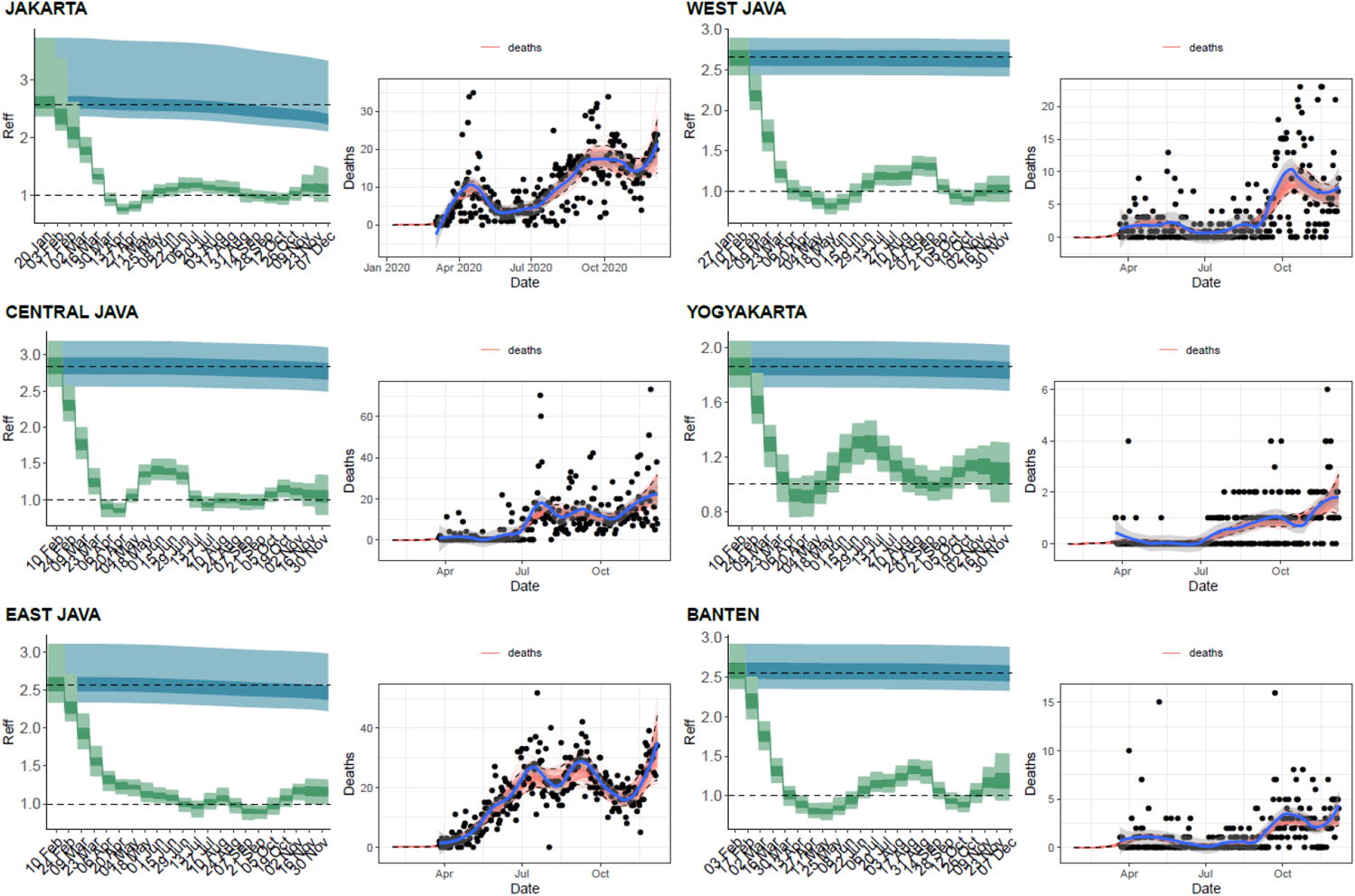
Model fits to reported deaths data and estimated ***R***_***c***_ values.

**Figure S10.**
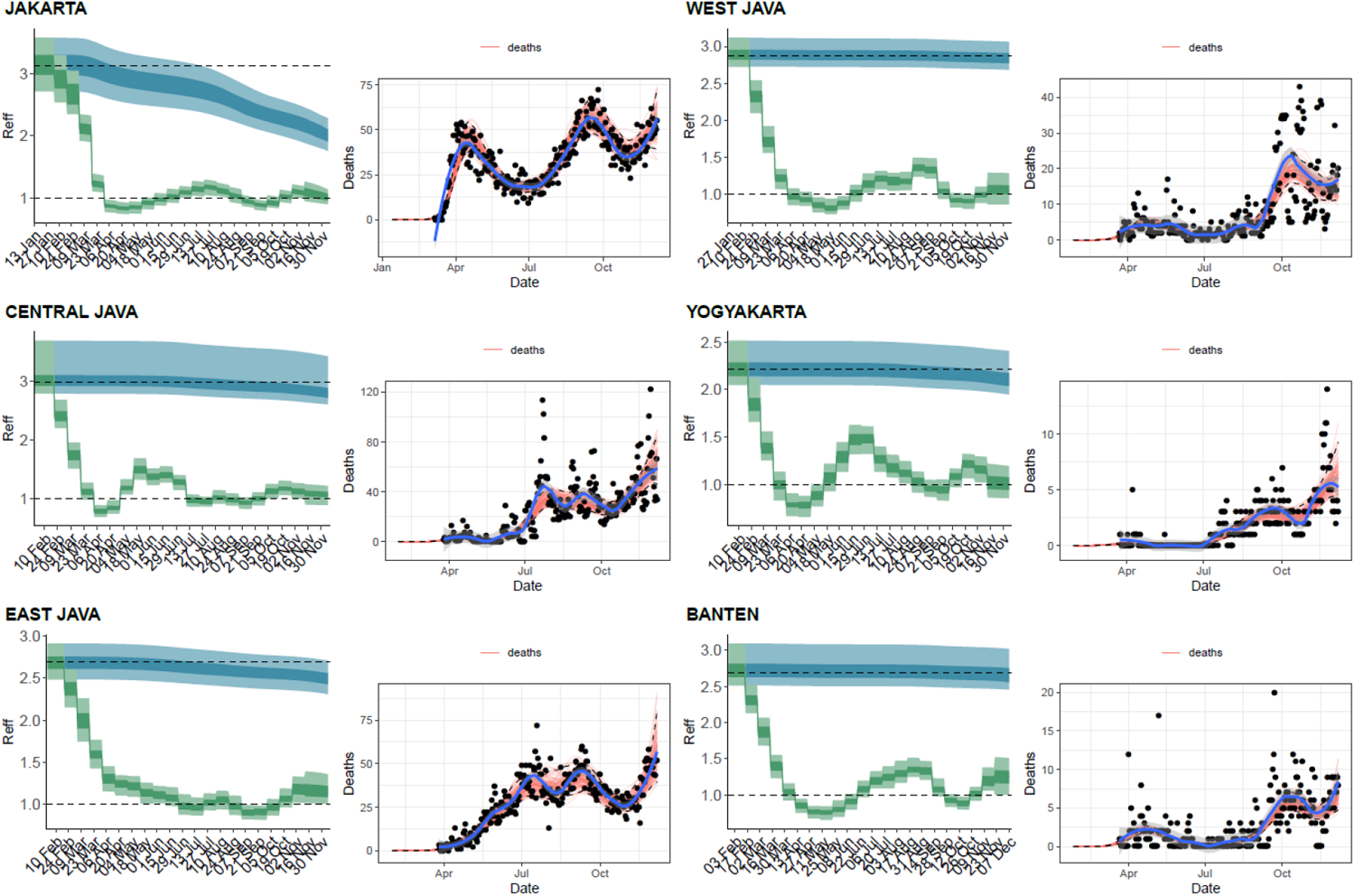
Model fits to suspected deaths data and estimated ***R***_***c***_ values.

**Figure S11.**
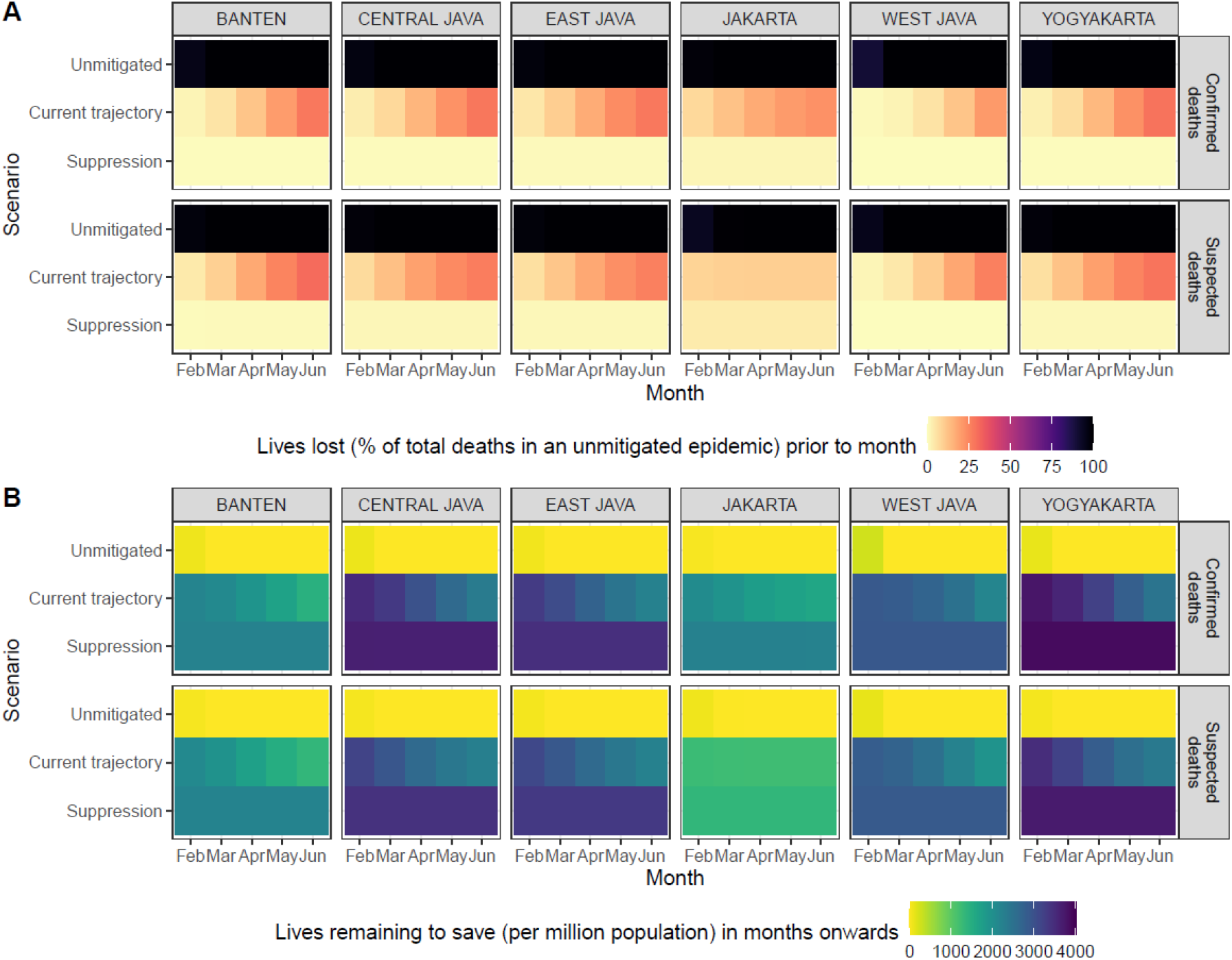
Additional analysis for projections of the number of lives lost and remaining to save (compared to Figure 6 in the main text), based on the assumptions of a higher reproduction number when ‘returning to normal’ and the epidemic is unmitigated (***R***_***c***_ = **3. 00**). A) Projected percentage of lives lost (compared to total deaths from an unmitigated epidemic scenario) prior to the start of each month from February to June 2021, based on each simulation scenario and model fitted to confirmed or suspected deaths in each province in Java; B) Projected number of lives remaining to be saved (or deaths that can still be averted) per million population after the start of each month from February to June 2021, based on each simulation scenario and model fitted to confirmed or suspected deaths in each province in Java.

**Figure S12.**
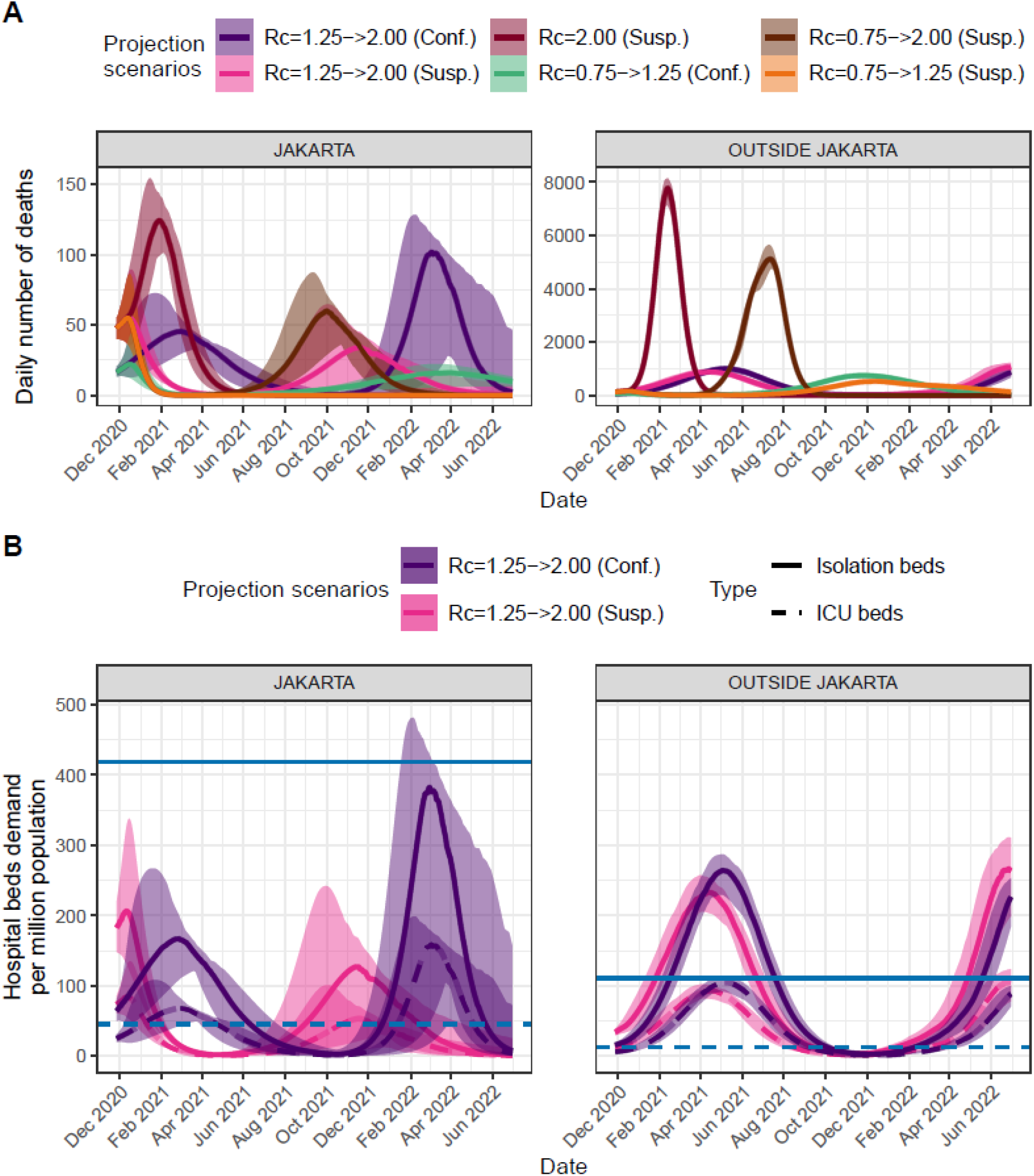
A) Projections of daily number of deaths due to COVID-19 based on six different transmission scenarios. B) Healthcare demand projections in the form of isolation beds and ICU beds demands assuming the current level of transmission to continue in the future (with easing of control measures after the transmission reached a low-level following the first peak in the graph). Projections are coloured according to whether they are based upon confirmed or suspected deaths to date and by projected ***R***_***c***_ (with ***R***_***c***_ = ***x*** → ***y*** representing ***R***_***c***_ = ***x*** for immediate future and ***R***_***c***_ = ***y***, the level it returns to when burden falls below 7 deaths per week). Healthcare capacities are based on the current numbers of dedicated COVID-19 isolation beds and ICUs^7^, not reflecting the total number of beds and ICUs in each province.

**Figure S13.**
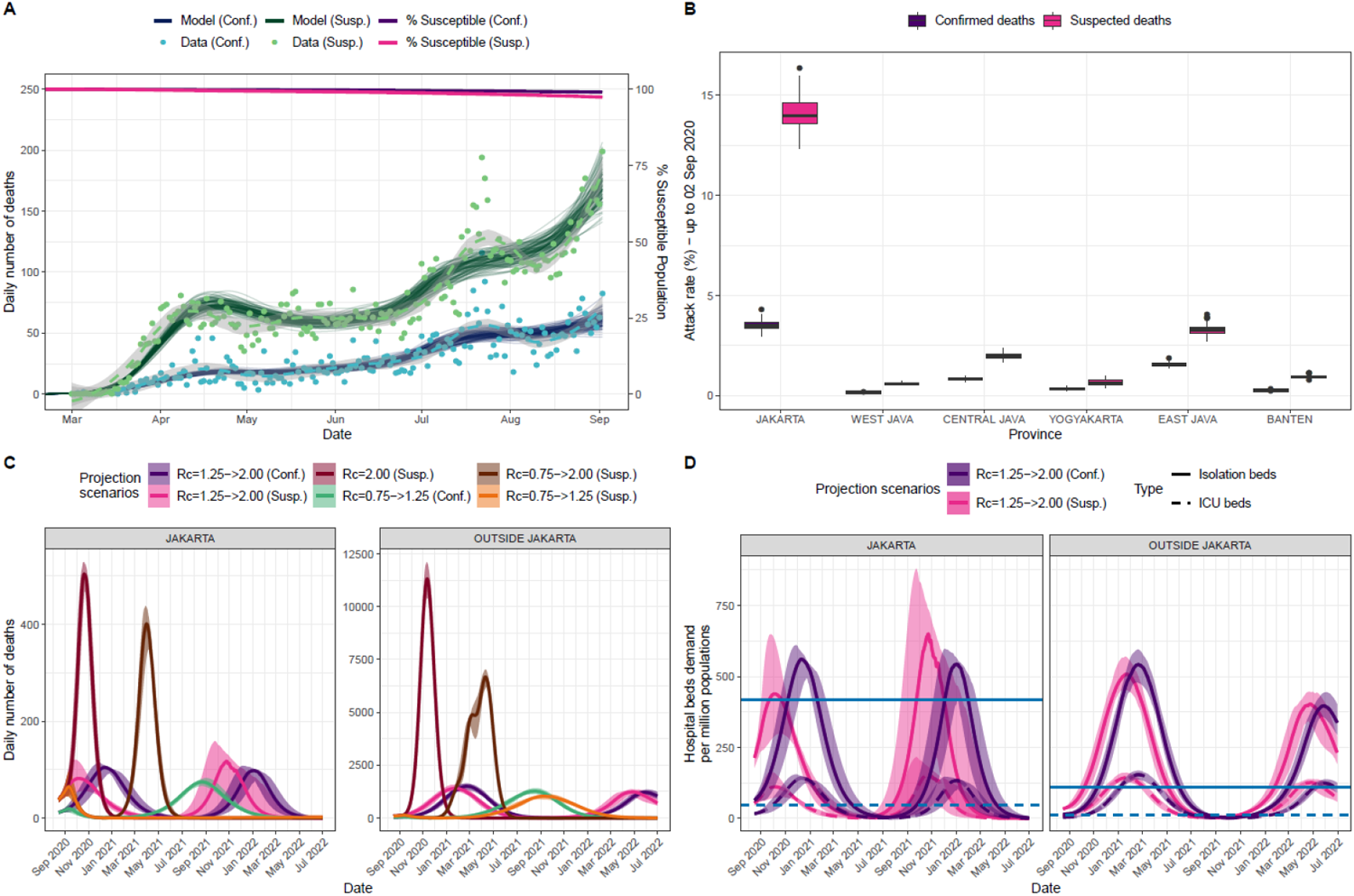
A) Model fitting to confirmed and suspected (both confirmed and probable) COVID-19 related deaths and inferred population susceptibility in Java; Green and blue dots show data on reported and suspected respectively (where suspected includes augmented estimate of probably deaths in provinces outside Jakarta pre-May 13th), with associated median (lines) and 95% CrI (shaded areas) of model fits. B) Estimated province-level attack rates (cumulative proportion infected) based on confirmed (purple) and suspected COVID-19 related deaths. C) Projections of daily number of deaths due to COVID-19 based on four different transmissibility scenarios. D) Healthcare demand projections in the form of isolation beds and ICU beds demands assuming the current level of transmission to continue in the future (with easing of control measures after the transmission reached a low-level following the first peak in the graph). Projections are coloured according to whether they are based upon confirmed or suspected deaths to date and by projected *R*_*c*_ (with *R*_*c*_ = *x*−> *y* representing *R*_*c*_ = *x* for immediate future and *R*_*c*_ = *y*, the level it returns to when burden falls below 7 deaths per week). Healthcare capacities are based on the current numbers of dedicated COVID-19 isolation beds and ICUs,^7^ not reflecting the total number of beds and ICUs in each province. The figure was taken from Djaafara et. al.^20^ (Figure 5).

**Figure S14.**
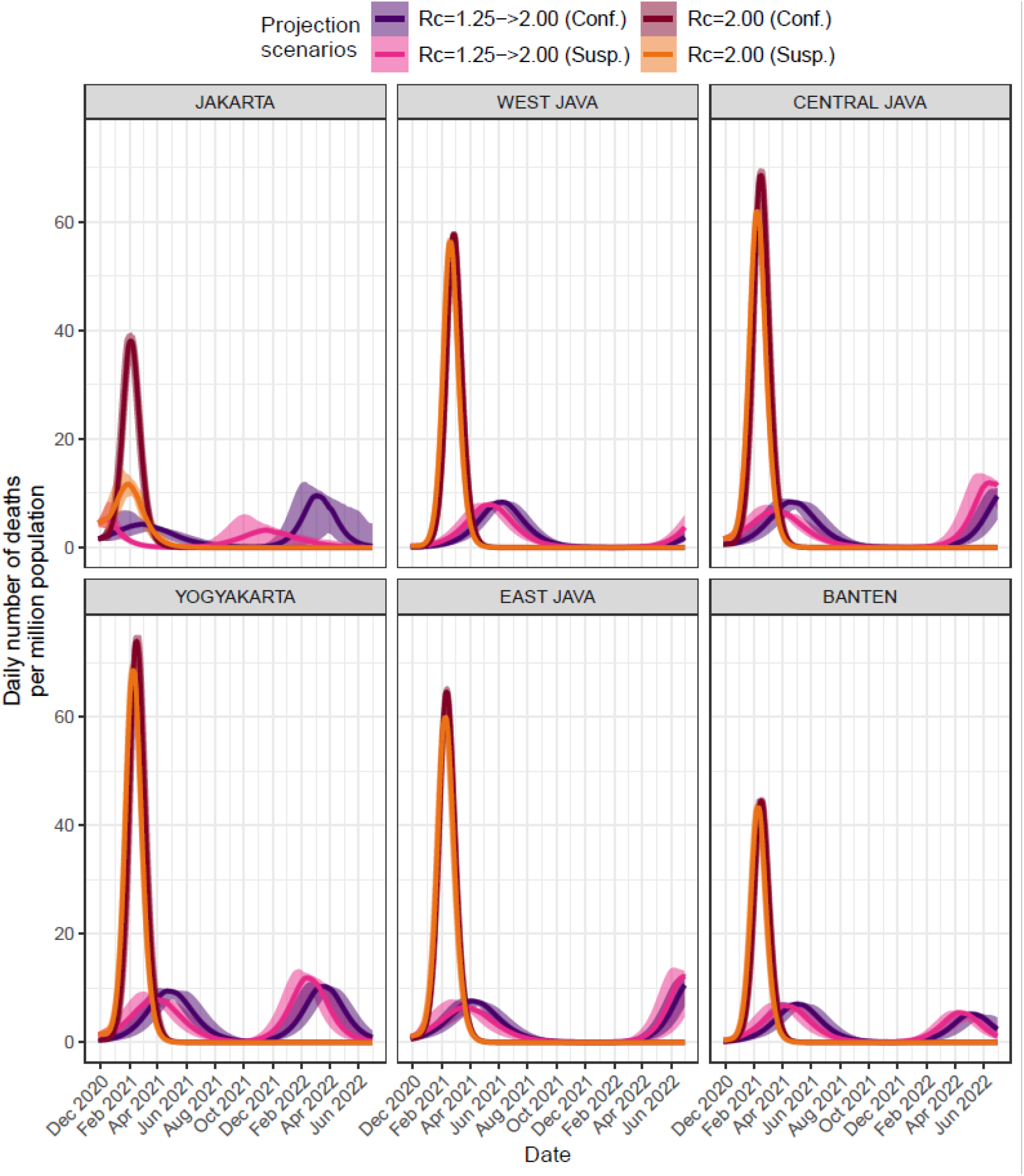
Projections of daily number of deaths due to COVID-19 based on four different transmission scenarios as shown in Figure 5C but showing all provinces in Java.

**Figure S15.**
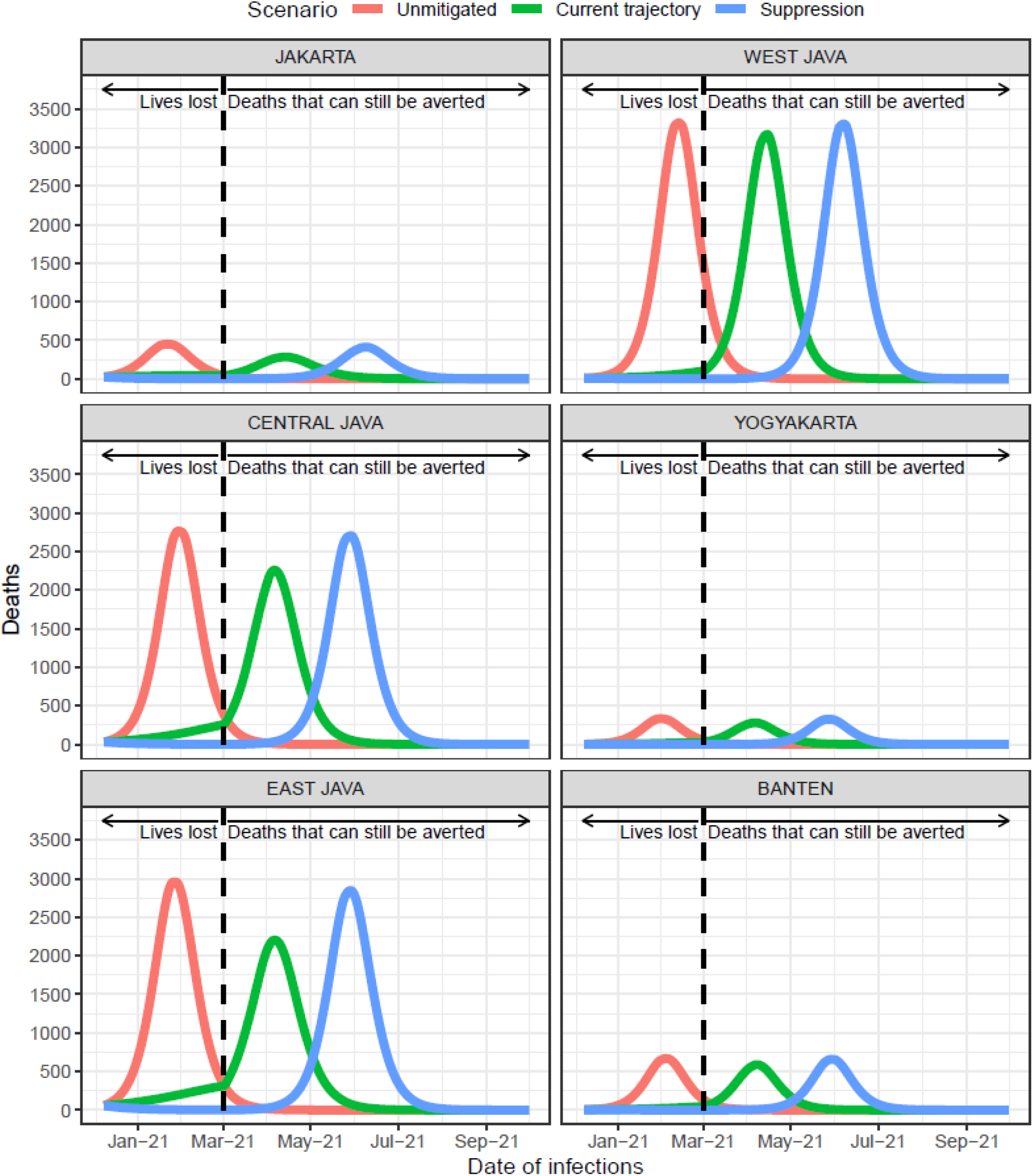
Illustrations of future scenario projections based on models fitted to confirmed COVID-19 deaths in each province in Java which subsequently ‘returning to normal’ on 1^st^ March 2021. The Jakarta trajectories are similar to what are shown in Figure 6A.

**Table S6.** Estimated attack rate in each province in Java island based on models fitted to confirmed or suspected deaths data on 2^nd^ September 2020 and 7^th^ December 2020.

| Province | Deaths data type | Estimation date | Attack rate in percentage (95% CrI) |
| --- | --- | --- | --- |
| JAKARTA | Confirmed deaths | 02-Sep | 4.61 (4.22-5.2) |
| JAKARTA | Confirmed deaths | 07-Dec | 9.75 (8.74-11.49) |
| JAKARTA | Suspected deaths | 02-Sep | 18.43 (17.04-19.66) |
| JAKARTA | Suspected deaths | 07-Dec | 32.48 (30.09-34.63) |
| WEST JAVA | Confirmed deaths | 02-Sep | 0.2 (0.17-0.24) |
| WEST JAVA | Confirmed deaths | 07-Dec | 0.66 (0.54-0.75) |
| WEST JAVA | Suspected deaths | 02-Sep | 0.41 (0.36-0.49) |
| WEST JAVA | Suspected deaths | 07-Dec | 1.42 (1.25-1.67) |
| CENTRAL JAVA | Confirmed deaths | 02-Sep | 0.97 (0.85-1.13) |
| CENTRAL JAVA | Confirmed deaths | 07-Dec | 2.3 (1.96-2.81) |
| CENTRAL JAVA | Suspected deaths | 02-Sep | 2.41 (2.05-2.88) |
| CENTRAL JAVA | Suspected deaths | 07-Dec | 5.96 (5.1-6.98) |
| YOGYAKARTA | Confirmed deaths | 02-Sep | 0.41 (0.33-0.49) |
| YOGYAKARTA | Confirmed deaths | 07-Dec | 1.39 (1.05-1.68) |
| YOGYAKARTA | Suspected deaths | 02-Sep | 1.12 (0.92-1.37) |
| YOGYAKARTA | Suspected deaths | 07-Dec | 4.09 (3.33-5.24) |
| EAST JAVA | Confirmed deaths | 02-Sep | 2.13 (1.89-2.33) |
| EAST JAVA | Confirmed deaths | 07-Dec | 3.85 (3.39-4.4) |
| EAST JAVA | Suspected deaths | 02-Sep | 3.35 (2.89-3.83) |
| EAST JAVA | Suspected deaths | 07-Dec | 6.17 (5.46-7.38) |
| BANTEN | Confirmed deaths | 02-Sep | 0.34 (0.29-0.41) |
| BANTEN | Confirmed deaths | 07-Dec | 1.08 (0.9-1.34) |
| BANTEN | Suspected deaths | 02-Sep | 0.59 (0.5-0.68) |
| BANTEN | Suspected deaths | 07-Dec | 2.09 (1.75-2.51) |

